# Comparative Diagnostic Evaluation of Real-Time PCR and Culture for Detecting Pathogens in Podiatric Wound Infections

**DOI:** 10.1101/2025.08.26.25334225

**Authors:** Mehdi Dehghani, Hans Norouzi, Shabnam Dehghan, Michelle L. Spruill, Mahmoud Goodarzi, Keagan H. Lee, Howard L. Martin

## Abstract

**Background:** Culture-based wound infection diagnostics have well-recognized limitations in detecting fastidious, anaerobic, or polymicrobial pathogens. This study compared the performance of a PCR wound panel with traditional culture.

**Methods:** Dual-swab specimens from 93 clinical wound cases underwent both culture and PCR testing. Analyses included organism-level concordance, Gram stain correlation, and resistance gene detection. Logistic regression, Receiver Operating Characteristic (ROC), and latent class analysis (LCA) were used to evaluate diagnostic performance.

**Results:** Under the conventional (culture-referenced) framework, PCR sensitivity was 98.3% and specificity was 73.5%. LCA (model M3) estimated PCR sensitivity at 95.6% and specificity at 91%. PCR detected 110 clinically significant pathogens missed or ambiguously reported (∼ 30%) by culture. A logistic model incorporating *16S rRNA* Ct values and Gram morphology found that 73.8% of PCR-only detections had infection probabilities >75%. Applying these probabilities and resistance gene codetection data in a symmetric framework improved PCR specificity from 45% to 86% with 95% sensitivity, while culture specificity remained high but suffered notable underdetection of known clinically significant pathogens. Many PCR-only detections, especially in polymicrobial wounds, were supported by high bacterial burden, Gram concordance, and resistance markers. For three key pathogens (*S. aureus, P. aeruginosa, S. agalactiae*), analysis showed PCR specificity ≥95% when compared to culture, suggesting that culture underdetection may partly reflect colony selection and prioritization during workup.

**Conclusion:** These results underscore the known limitations of culture in resolving polymicrobial podiatric wound infections and highlight PCR’s role in providing significantly faster and more comprehensive organism-level detection to guide clinical decisions.

## Introduction

Wound infections pose a serious clinical challenge, increasing patient morbidity, hospitalization rates, and healthcare costs, especially for patients with surgical wounds, diabetic foot ulcers, pressure ulcers, and other chronic ulcers (1, 2). Detecting microorganisms accurately and in a timely manner is crucial for ensuring effective antimicrobial treatment and minimizing severe complications, such as osteomyelitis or amputation, particularly in patients with diabetic foot ulcers (3, 4).

Traditional culture, long considered the gold standard for wound infection diagnosis, relies on live microorganisms, appropriate growth conditions, and trained microbiologists, which can introduce an element of subjectivity (5). These limitations may decrease sensitivity, especially in polymicrobial, anaerobic, or antibiotic-treated wounds. Multiple studies have documented the drawbacks of culture-based diagnostics in diabetic foot infections, emphasizing the need for more sensitive and specific methods to improve patient outcomes (6, 7).

Recent advancements in molecular techniques have provided new tools for microbial detection. PCR has shown promise, as it can identify infectious agents faster and more accurately than traditional culture methods (8, 9). Molecular methods can simultaneously identify a broad range of microorganisms, including anaerobes, fastidious bacterial species, and antibiotic-resistant pathogens, giving clinicians precise and actionable diagnostic information (10, 11). However, PCR methods can detect DNA from non-viable organisms or normal flora, raising concerns about false positives.

In this study, we compared the diagnostic performance of a comprehensive PCR panel (BioExcel Diagnostics) with that of traditional cultures performed by a commercial reference laboratory. Using statistical methods such as logistic regression, receiver operating characteristic (ROC) curves, and latent class analysis (LCA) (12), we objectively evaluated the diagnostic accuracy of PCR relative to traditional culture. Our findings address critical gaps in podiatric wound infection diagnostics and provide evidence to guide clinical practice.

## Materials and Methods

### Study Design and Specimen Collection

As part of a quality assurance program, we compared traditional culture results (performed by a reputable commercial laboratory) with those from an analytically validated comprehensive PCR panel (BioExcel Diagnostics). We reviewed 107 wound infection cases collected between January 2024 and January 2025, in which providers had requested both culture and PCR testing. Specimens primarily included diabetic foot ulcers, pressure ulcers, surgical sites, and other soft tissue infections suspected of microbial involvement. Samples were predominantly collected using swabs, with 49 cases classified as unspecified swabs (45.8%) and 37 as ulcer-specific swabs (34.6%). Less common specimen types included shave biopsies (5.6%), abscess swabs (4.7%), and bone tissue (1.9%). The remaining categories consist of deep tissue swabs, aspirates, excisional biopsies, and other rare types. This dual-swab approach minimized sampling variability and facilitated an unbiased, parallel comparison between molecular PCR testing and conventional culture-based methods (10).

### Culture-Based Testing and Gram Stain Assessment

Culture and antimicrobial susceptibility testing (AST) were performed by a reputable commercial reference laboratory. Gram stain results were reviewed for morphological features, including Gram-positive cocci (GPC), Gram-negative rods (GNR), epithelial cells, and white blood cells (WBC). Specific methodological details were unavailable; therefore, our analysis relied solely on the final culture reports for each case.

### DNA Extraction and PCR Testing

Nucleic acid extraction was conducted using the MagMAX™ Microbiome Ultra Nucleic Acid Isolation Kit (Thermo Fisher Scientific). Mechanical bead-based lysis was performed using the Omni Bead Ruptor Elite 24 to effectively disrupt bacterial cell walls for both Gram-positive and Gram-negative organisms. Real-time PCR testing was performed using the nanoscale SmartChip Real-Time PCR System (Takara Bio). We employed widely used TaqMan assays (Thermo Fisher Scientific) targeting numerous clinically relevant bacterial and fungal pathogens, as well as key antibiotic resistance genes (ABR). Each PCR run included internal controls for extraction efficiency, amplification performance, and contamination, as well as appropriate positive and negative controls.

### Case Inclusion and Diagnostic Classification

Of the 107 wound infection cases initially reviewed, 93 cases with complete paired data (PCR and culture results) were included in the final analysis. Out of 107 initial cases, two cases were excluded due to ambiguous culture results listing only "mixed flora," and twelve were removed due to incomplete or inconclusive culture data. Diagnostic classification was performed using a conventional reference framework, in which culture served as the reference. Cases were classified as true positive (TP) if PCR and culture identified at least one matched clinically significant organism. Cases were considered false positives (FP) if PCR detected one or more clinically pathogenic microorganisms that were not recovered by culture, and there were no matched organisms between the two methods. False negatives (FN) were assigned to cases when culture detected a pathogenic organism that PCR did not detect. True negatives (TN) were cases where neither method detected any clinically significant pathogen. To enable a like-for-like comparison between PCR and culture, organisms typically regarded as commensals (e.g., coagulase-negative staphylococci) or fastidious anaerobes (e.g., *Finegoldia magna*, *Actinotignum schaalii*, *Peptoniphilus spp*.) were excluded from classification.

### Statistical Analyses

Diagnostic performance metrics, including sensitivity, specificity, positive predictive value (PPV), negative predictive value (NPV), accuracy, and F1 score, were calculated. Discordance between PCR and culture was statistically evaluated using McNamar’s exact test. Additionally, we evaluated the predictive power of pan-bacterial burden (16S rRNA Ct values), organism-specific Ct thresholds, and Gram stain morphology in determining true infection status using logistic regression models. We used Receiver Operating Characteristic (ROC) curves to assess model performance. Latent class analysis (LCA) objectively estimated diagnostic accuracy without assuming a definitive reference standard (12). Latent class analyses used two classes (infected vs. not infected) and assumed local independence. To avoid non-identification inherent to two-indicator models, we treated the two-indicator model (M1: PCR + culture) as descriptive only and based inference on the three-indicator model (M3: PCR + culture + Gram). Models were fitted by EM with 100 random starts. Parameter uncertainty was quantified via a 300-replicate parametric bootstrap (seed = 2025); the LCA_broad_panel_pipeline.py would be available upon request.

### Predictive Modeling

A morphology-informed logistic regression model estimated the infection probability/likelihood for PCR-detected organisms. Predictors included organism-specific Ct values, total bacterial load (as measured by *16S rRNA* Ct), and Gram stain morphology scores. The model was trained exclusively on confirmed endpoints (true positives and negatives) and validated using internal metrics such as ROC analysis and calibration curves. Infection probabilities were assigned to PCR-only organism detections, providing continuous likelihood scores validated by simulation analysis and concordance with resistance genes. The model was trained exclusively on confirmed biological endpoints, organisms detected by both PCR and culture, and was validated using cross-validation and calibration methods (12–16). For logistic regression using *16S rRNA* Ct + Gram_Score_Sum, calibration was done by Platt scaling (sigmoid) with 5-fold out-of-fold (OOF) evaluation. OOF metrics are as follows: intercept −0.053 (95% CI −0.803–0.698), slope 1.051 (0.548–1.555), Brier score 0.143, ECE(10) 0.085, HL p=0.059. Calibration was close to ideal, and tight in the 0.8–1.0 region.

### Power Analysis

A power analysis confirmed that the evaluable sample size (n = 93) provided greater than 90% power to detect clinically meaningful differences in specificity (≥20% improvement) at a significance level of *α* = 0.05, ensuring reliable and robust comparative evaluations between PCR and culture performances.

## Results

### Descriptive overview of the study cohort

In this retrospective study, an initial review of electronic records identified 107 cases of wound infection for which the corresponding healthcare provider requested both PCR and traditional culture simultaneously. As shown in **Figure S1**, after excluding 2 cases with mixed or unidentified culture results and 12 cases deemed inconclusive due to limitations in colony differentiation for aerobic and anaerobic cultures, the final evaluable set consisted of 93 cases with paired PCR and culture data, 59 culture-positive and 34 culture-negative. Notably, in the 12 cases where culture results were inconclusive, PCR still provided a complete organism-level profile. Specimens were predominantly swabs, 49 unspecified (45.8%) and 37 ulcer-targeted (34.6%), with fewer shave biopsies (5.6%), abscess swabs (4.7%), and bone and deep-to-bone samples (each 1.9%); several additional types were each <1% (e.g., aspirate, blister swab) (**Supplementary Table S1**). Demographically, 56.0% of cases were from men, 38.3% from women (5.5% unspecified); the median age was 67 years (range 13–90), with the distribution skewed toward older ages. Most specimens originated from podiatric sites (∼90%).

PCR reports were available in a median of 20 h (IQR 4.5 h) versus 120 h for culture (IQR 0 h), a >4-day advantage (paired Wilcoxon p = 2.57×10⁻¹⁹). The culture lab’s IQR = 0 indicates batch/scheduled release with little mid-distribution variability; PCR varied modestly around a ∼20-hour median, reflecting more responsive specimen processing. **(Supplementary Table S2; Figure S2).**

Across the evaluable 93 cases, PCR detected a broad range of pathogens (**Supplementary Table S3**), including common aerobes (e.g., *Staphylococcus aureus*, *Pseudomonas aeruginosa*, *Streptococcus agalactiae*) and fastidious/anaerobic taxa (e.g., *Finegoldia magna*, *Bacteroides fragilis*) that are often under-recovered by culture (10, 17). Notably, *Staphylococcus aureus* remains the most commonly detected organism (1). Culture detections for the cohort are summarized in **Supplementary Table S4** (e.g., *Staphylococcus aureus*; 36/93, *Pseudomonas aeruginosa;* 15/93, *Streptococcus agalactiae;* 5/93).

### Performance evaluation under the conventional, culture-referenced classification

Case-level comparison between the culture and PCR outcomes is shown in **Figure 1**: 17 fully concordant TPs, 25 partial TPs (PCR detected ≥1 additional organism), 16 TPs with a culture flag of unidentified additional organisms (TP-UAO), 9 FPs (PCR+/culture−), 1 FN (PCR−/culture+), and 25 TNs (n = 93). From these counts (58 TP, 9 FP, 1 FN, 25 TN), PCR achieved a sensitivity of 98.3% (58/59) and specificity of 73.5% (25/34) compared to culture. The positive predictive value (PPV) was 86.8%, the negative predictive value (NPV) was 96.3%, and the overall diagnostic accuracy and F1 score were 89.3% and 92.1%, respectively (**Table 1**). These metrics were calculated after excluding organisms considered normal flora or fastidious (e.g., *Coagulase-Negative Staphylococci*, *Actinotignum schaalii*) to ensure a direct, like-for-like comparison with culture.

**Figure 1.**
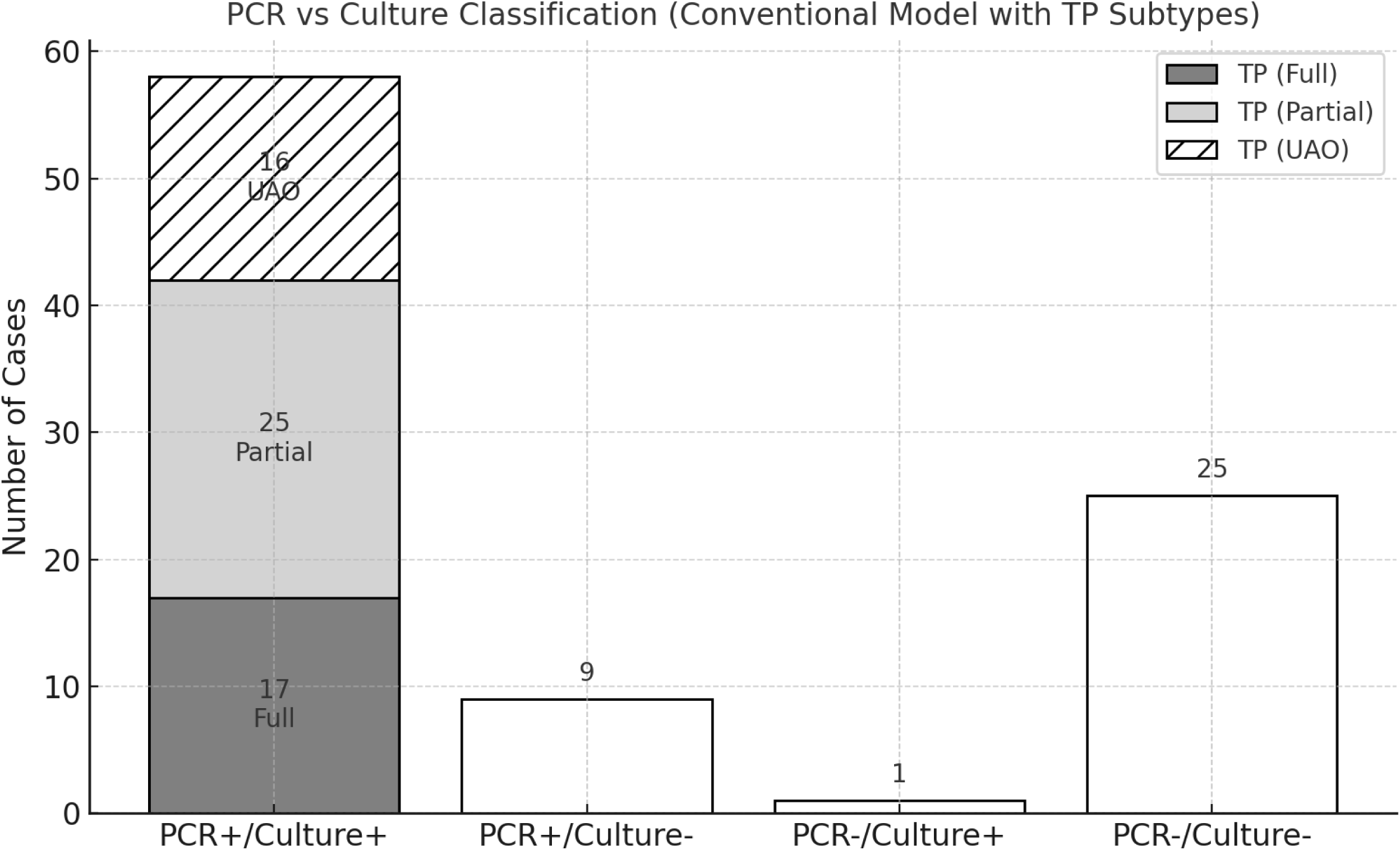
Case-level classification of wound infection using the conventional classification model (culture-referenced). Among 93 evaluable cases, 58 were PCR+/Culture+, including 17 fully concordant cases (TP Fully concordant), 25 partially concordant cases with additional PCR-only organisms (TP Partial), and 16 cases with culture-reported “unidentified additional organisms” (TP UAO). The remaining cases included 9 PCR-positive/culture-negative (FP), 1 PCR-negative/culture-positive (FN), and 25 PCR-negative/culture-negative (TN).

**Table 1.**
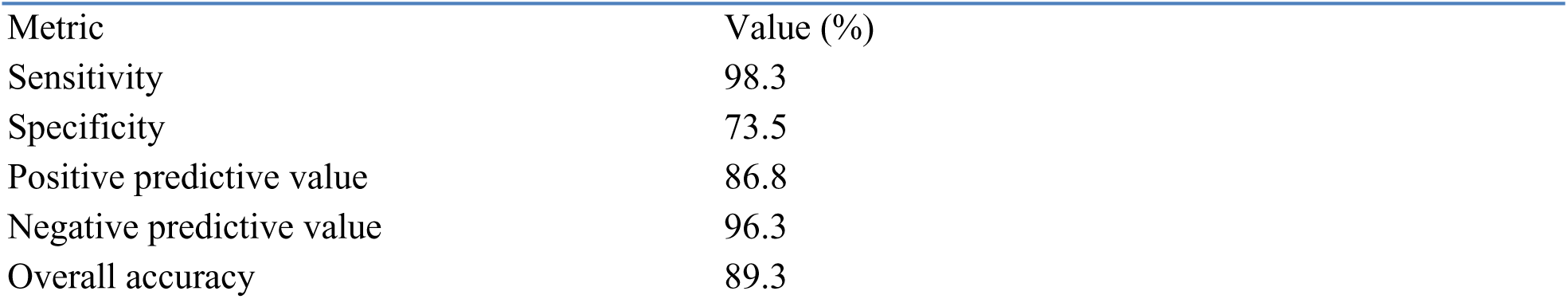
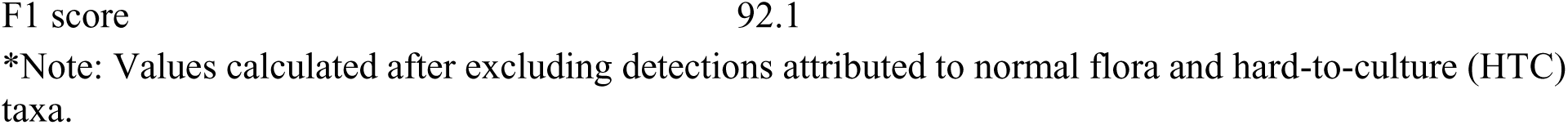
Diagnostic performance of PCR compared with culture using conventional classification model (n = 93 cases).

| Metric | Value (%) |
| --- | --- |
| Sensitivity | 98.3 |
| Specificity | 73.5 |
| Positive predictive value | 86.8 |
| Negative predictive value | 96.3 |
| Overall accuracy | 89.3 |

\*Note: Values calculated after excluding detections attributed to normal flora and hard-to-culture (HTC) taxa.

To further compare diagnostic yield, McNemar’s exact test was applied at both the case and organism levels. At the case level (**Figure 2A**), there were 9 PCR-only vs one culture-only case, showing significant asymmetry (p = 0.021). At the organism level, restricting to panel organisms (**Figure 2B**), 50 cases had PCR-only additional organisms versus three cases with culture-only detections (p = 5.5×10^-^¹²). Including all organisms (**Figure 2C**), i.e., counting culture-only detections outside the PCR panel, PCR-only detections still dominated (50 vs 12, p = 1.2×10⁻⁶). Across these partially concordant or discordant cases, PCR identified 110 additional organisms (median 2; mean 2.2 per case); 64% of cases had >1 additional organism (**Supplementary Table S5**), consistent with molecular studies showing under-recovery of mixed communities by culture (13). Thirty-eight out of the 110 PCR-only organisms were from 15 TP cases that contained the UAO flag.

**Figure 2.**
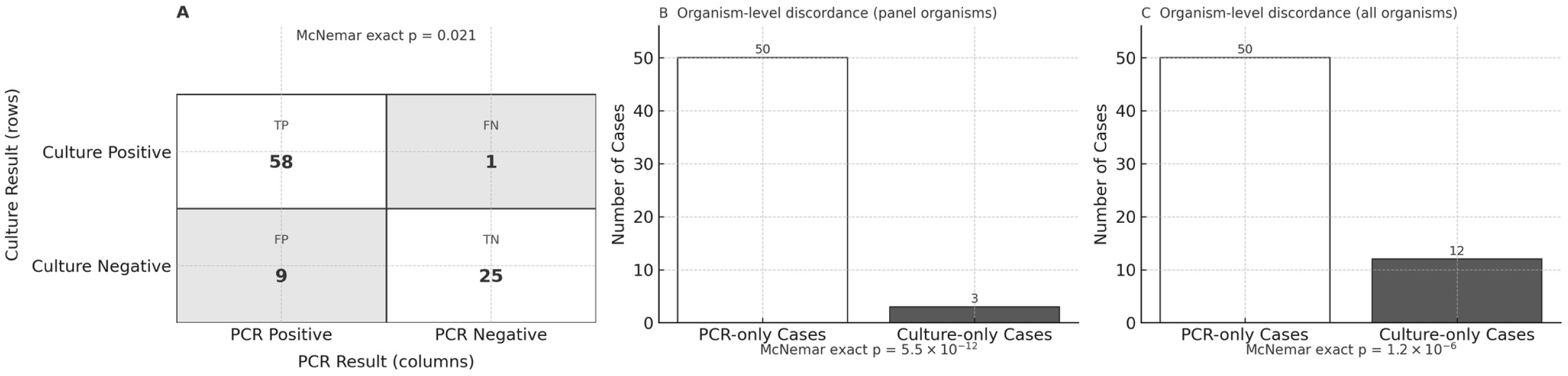
Directional discordance between PCR and culture results at the case and organism levels. (A) Case-level confusion matrix (n = 93): PCR positive / culture positive = 58; PCR negative / culture positive (FN) = 1; PCR positive / culture negative (FP) = 9; PCR negative / culture negative = 25; McNemar exact p = 0.021(statistically significant). (B) Organism-level discordance limited to panel organisms (targets covered by the PCR panel): 50 cases with PCR-only calls vs 3 cases with culture-only detections; McNemar exact p = 5.5×10^−12^ (statistically significant) (C) Organism-level discordance including off-panel culture detections (all organisms): 50 cases with PCR-only calls vs 12 cases with culture-only detection; McNemar exact p = 1.2×10^−6^ (statistically significant).

Overall, this organism-level discrepancy between the studied PCR panel and the culture results from the reference laboratory raises an important question: Are these additional organisms detected by PCR incidental, or do they represent clinically significant parts of the infection that culture failed to identify?

### Organism-level performance evaluation for key pathogens (culture-referenced)

To stress-test potential nonspecific amplification or amplification from non-viable organisms, we first reviewed 28 cases (28-subset) in which the culture reports explicitly ruled out the presence of Pseudomonas aeruginosa, Staphylococcus aureus, and Streptococcus agalactiae (**Supplementary Table S6**). In this cohort, PCR yielded no detections for *P. aeruginosa* and *S. agalactiae* (100% specificity for each) and only one detection for *S. aureus*; aggregated across 84 organism-events, specificity was 98.8% (83/84) (binomial p = 0.0015 vs a ≤90% null; Bayesian mean 97.7%). Across the 93 evaluable cases, culture-referenced organism-level performance for these three key pathogens was high overall, with sensitivities 86.7–100% and specificities 94.0–99.0%; the exact per-organism performance characteristics are summarized in **Table 2**. To minimize repetition, we summarize the results for another specificity evaluation strategy here and provide details in **Table 2’s** footnote: in a subset of 77 cases (comprising all included cases except for TPs with "unidentified additional organisms" (UAO) comments), there were 6 PCR-positive results among 195 culture-negative organism events. Consequently, the overall specificity was calculated as 96.9% (189/195), with a binomial p-value of 0.00022. The Bayesian posterior mean specificity was estimated at 96.4%, with a 95% credible interval (CrI) ranging from 93.5% to 98.6%.

**Table 2.** Diagnostic Performance of PCR for S. *aureus, P. aeruginosa* and *S. agalactiae* (n = 93)

| Organisms | TP | FP | FN | TN | Sensitivity | Specificity | PPV | NPV | Accuracy | F1 Score |
| --- | --- | --- | --- | --- | --- | --- | --- | --- | --- | --- |
| *Staphylococcus aureus* | 35 | 3 | 1 | 54 | 0.97 | 0.95 | 0.92 | 0.98 | 0.96 | 0.95 |
| *Pseudomonas aeruginosa* | 13 | 1 | 2 | 77 | 0.87 | 0.99 | 0.93 | 0.97 | 0.97 | 0.90 |
| *Streptococcus agalactiae* * | 5 | 5 | 0 | 83 | 1.00 | 0.94 | 0.50 | 1.00 | 0.95 | 0.67 |

† 28case cohort: Culture explicitly ruled out the growth of *S. aureus*, *P. aeruginosa*, and *S. agalactiae* (see Suppl. Table S6). PCR: 0/28 for *P. aeruginosa* and *S. agalactiae*; 1/28 for *S. aureus*. Aggregated specificity 98.8% (83/84); binomial $p=0.0015$ (statistically significant); Bayesian mean 97.7%. [UAO = Unidentified Additional Organisms.
‡ 77case subset (no UAO comment in culture reports): 231 organism events (77×3). Of 195 culture-negative events, only 6 were PCR-positive, yielding a specificity of 96.9% (189/195); binomial $p=0.00022$ (statistically significant); Bayesian posterior mean 96.4% (95% Credible Interval: 93.5–98.6%); $P(\text{specificity}>0.90) = 99.98\%$ ; TP: True Positives; FP: False Positives; FN: False Negatives; TN: True Negatives; PPV: Positive Predictive Value; NPV: Negative Predictive Value.
Note: Three out of 5 cases labeled as FPs for *S. agalactiae* had a culture note indicating the presence of “Unidentified Additional Organisms” (UAO).

Together, these results argue against widespread false-positive PCR detection or amplification of non-viable cells for these key pathogens.

### Detection prevalence and culture under-recovery of *S. agalactiae*

By detection rates across 107 cases, PCR vs culture identified *S. aureus* in 35.5% vs 33.6%, *P. aeruginosa* in 13.1% vs 14.0%, and *S. agalactiae* in 10.3% vs 5.6%, respectively (**Table 3**). Among 11 PCR-positive *S. agalactiae* cases, six were culture-concordant and five were culture-negative. Example profiles include Case 71 (low-Ct S. *agalactiae* [Ct 19.2] co-detected with *A. baumannii* [Ct 19.9], *S. maltophilia* [Ct 19.2], *E. faecalis* [Ct 24.4], *S. aureus* [Ct 18.7], plus an *OXA-group* carbapenemase [Ct ∼19]), and Cases 92 and 104 (Ct 25.4 and 30.4 for *S. agalactiae*). Notably, 3/5 discordant reports (Cases 71, 92, 104) contained the UAO comments, consistent with the overall frequency of such notes (30.8%, 33/107), among all 107 cases. These patterns support plausible S. *agalactiae* under-recovery by culture in polymicrobial wounds (details in Supplementary Table S7).

**Table 3.** PCR and Culture detection rates for Staphylococcus aureus, Streptococcus agalactiae, and Pseudomonas aeruginosa.

| Organism | PCR Detection Count | PCR Detection Rate (%) | Culture Detection Count | Culture Detection Rate (%) |
| --- | --- | --- | --- | --- |
| *S. aureus* | 38 | 35.5 | 36 | 33.6 |
| *S. agalactiae* | 11 | 10.3 | 6 | 5.6 |
| *P. aeruginosa* | 14 | 13.1 | 15 | 14.0 |

\*Note: Detection rates are based on 107 total cases. PCR rates reflect detections above Ct, Gram stain, or resistance gene thresholds.

### *Enterococcus faecalis*: missed by culture, supported by PCR

PCR detected *Enterococcus faecalis* in 36/107 cases (33.6%), with Ct ≤ 26 in 73% and Gram-positive cocci on smear in 89%, yet none were reported as *E. faecalis* by culture in this cohort. In the 77-case high-confidence specimen subset (no TP-UAO), organism-specific Ct distributions for PCR-only *E. faecalis* (labeled FP under the culture-referenced framework; n = 21) were comparable to culture-confirmed *S. aureus* TPs (n = 23): median Ct 22.5 vs 19.2 (Mann–Whitney p = 0.0128), with overlapping ranges (17.9–30.7 vs 15.3–31.3; Kolmogorov–Smirnov p = 0.066; ∼80% overlap; Figure 3). These data indicate that PCR-only *E. faecalis* detections carry organism burden and Ct distribution patterns similar to the *S. aureus* detections, supporting biological plausibility rather than low-level noise. (Probability-based modeling for *Enterococci* is reported later; see **Table 4**.)

**Figure 3.**
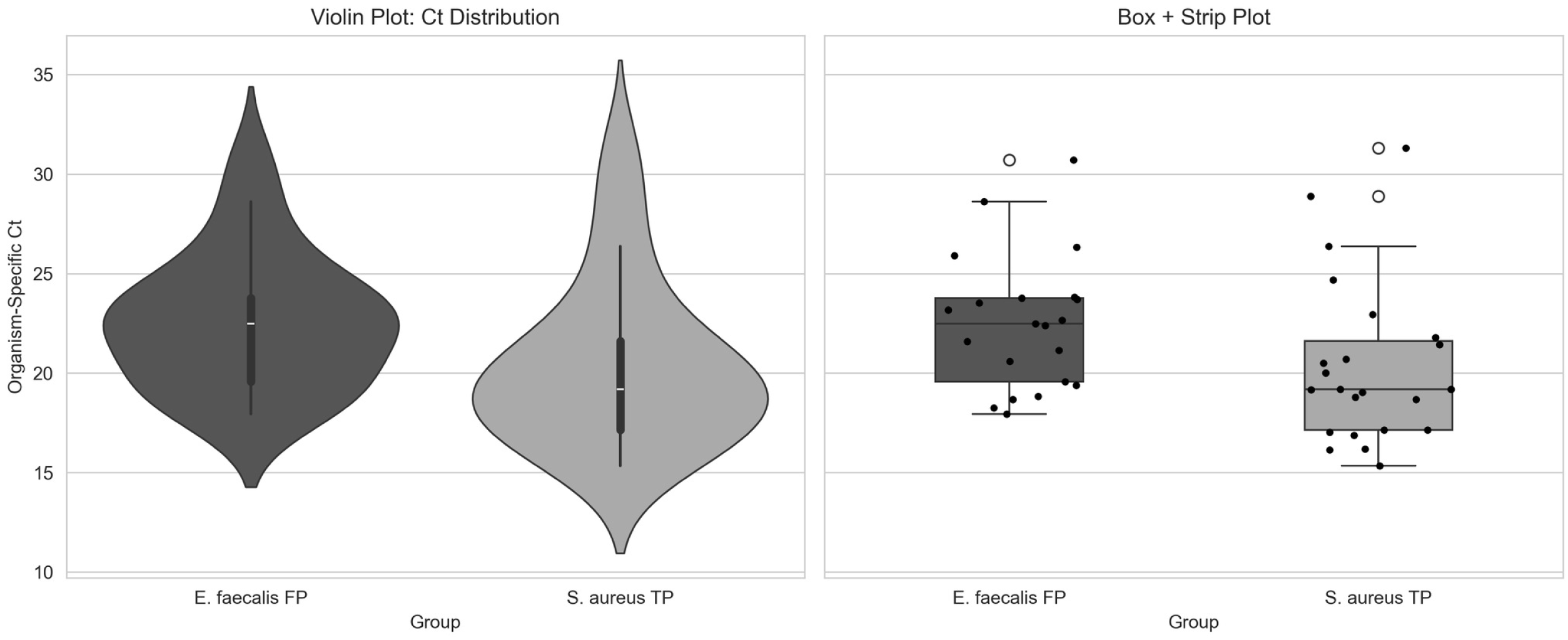
Comparison of Organism-Specific Ct Values Between *Enterococcus faecalis* False Positives and *Staphylococcus aureus* True Positives. Violin plot (left) and box + strip plot (right) show the distribution of *E. faecalis* Ct values classified as false positives (n = 21) and culture-matched *S. aureus* true positives (n = 23) in all included cases that had no UAO flag. While *E. faecalis* FPs had slightly higher median Ct values (22.5 vs. 19.2), both groups showed substantial overlap in distribution (80% Ct range overlap), and the Kolmogorov–Smirnov test did not detect a statistically significant difference in shape (p = 0.066; not significant). These findings support the biological plausibility of *E. faecalis* PCR-only detections and suggest they represent clinically relevant signals rather than background noise or nonviable DNA.

**Table 4.** Predicted Infection Probabilities for *Enterococcus* spp. PCR Detections. summarizes predicted infection probabilities for *E. faecalis* and *Enterococcus faecium* PCR detections,

| Case No. | Organism | Ct | GPC | Calibrated Prob. | $\geq 80\%$ calls | Possible ABR genes | ABR Cts |
| --- | --- | --- | --- | --- | --- | --- | --- |
| 1 | *E. faecium* | 31.3 | Low | 0.717 | 0 | *van* | 27.7 |
| 7 | *E. faecalis* | 22.6 | Absent | 0.807 | 1 | *erm(A)/erm(B); tet(M)* | 21.6; 22.5 |
| 8 | *E. faecalis* | 30.7 | Absent | 0.285 | 0 | *erm(A)/erm(B)* | 28.6 |
| 12 | *E. faecalis* | 18.8 | Low | 0.958 | 1 | *erm(A)/erm(B); tet(M)* | 17.74; 18.16 |
| 13 | *E. faecalis* | 28.6 | High | 0.651 | 0 | *erm(A)/erm(B); tet(M)* | 27.7; 27.96 |
| 15 | *E. faecalis* | 17.9 | High | 0.980 | 1 | *erm(A)/erm(B); tet(M)* | 16.9; 17.4 |
| 18 | *E. faecalis* | 19.4 | High | 0.970 | 1 |  |  |
| 19 | *E. faecalis* | 23.8 | High | 0.940 | 1 | *erm(A)/erm(B)* | 19.2 |
| 22 | *E. faecalis* | 23.2 | High | 0.942 | 1 | *erm(A)/erm(B); tet(M)* | 23.16; 22.6 |
| 25 | *E. faecalis* | 20.7 | High | 0.960 | 1 | *erm(A)/erm(B); tet(M)* | 20.3; 20.5 |
| 28 | *E. faecalis* | 23.8 | High | 0.918 | 1 | *tet(M)* | 23.8 |
| 29 | *E. faecalis* | 20.6 | High | 0.966 | 1 | *erm(A)/erm(B); van* | 30.7; 30.8 |
| 41 | *E. faecalis* | 19.6 | High | 0.967 | 1 | *erm(A)/erm(B); tet(M)* | 18.9; 19.4 |
| 47 | *E. faecalis* | 22.4 | Low | 0.936 | 1 | *tet(M); erm(A)/erm(B)* | 23.4; 29.93 |
| 48 | *E. faecalis* | 23.52 | High | 0.920 | 1 | *erm(A)/erm(B); van* | 24.7; 31.5 |
| 52 | *E. faecalis* | 23.5 | Low | 0.926 | 1 | *tet(M)* | 24.4 |
| 55 | *E. faecium* | 27.2 | High | 0.881 | 1 | Inconclusive | Inconclusive |
| 55 | *E. faecalis* | 18.3 | High | 0.979 | 1 | Inconclusive | Inconclusive |
| 58 | *E. faecalis* | 27.9 | Medium | 0.819 | 1 | *erm(A)/erm(B)* | 23.26 |
| 59 | *E. faecium* | 30.9 | Low | 0.663 | 0 | *van* | 28.05 |
| 61 | *E. faecalis* | 21.6 | High | 0.891 | 1 | *tet(M)* | 21.2 |
| 67 | *E. faecalis* | 26.32 | High | 0.885 | 1 | *tet(M)* | 25.1 |
| 69 | *E. faecalis* | 21.1 | High | 0.904 | 1 | *erm(A)/erm(B); tet(M)* | 20.6; 21.15 |
| 70 | *E. faecalis* | 22.5 | Medium | 0.923 | 1 | *tet(M)* | 22.1 |
| 71 | *E. faecalis* | 24.4 | High | 0.924 | 1 | *erm(A)/erm(B); tet(M)* | 20.7; 18.4 |
| 78 | *E. faecalis* | 18 | High | 0.971 | 1 | *erm(A)/erm(B); tet(M)* | 26.8; 26.5 |
| 83 | *E. faecalis* | 23.7 | High | 0.829 | 1 | Inconclusive | Inconclusive |
| 93 | *E. faecalis* | 27.2 | Low | 0.850 | 1 | Inconclusive | Inconclusive |
| 95 | *E. faecalis* | 26.6 | High | 0.881 | 1 | *erm(A)/erm(B); tet(M)* | 26.6; 25.5 |
| 98 | *E. faecalis* | 23.1 | Medium | 0.814 | 1 | *erm(A)/erm(B)* | 23.3; 22.6 |
| 101 | *E. faecalis* | 18.7 | Absent | 0.973 | 1 | *tet(M)* | 18.5 |
| 104 | *E. faecalis* | 30.9 | Absent | 0.742 | 0 | *tet(M)* | 30.7 |
| 105 | *E. faecalis* | 25.9 | High | 0.626 | 0 | *tet(M)* | 25.9 |

### PCR-Only GNRs are backed by Gram stain and ABR markers

Among 39 Gram-stain GNR–positive cases (of n = 93; included cases), PCR and culture were concordant in 17; 13 were PCR-positive/culture-negative; 1 was culture-positive/PCR-negative; and eight were double-negative **(Figure 4).** Notably, 7/13 PCR-positive/culture-negative cases also contained at least one organism matched by both methods, yet the culture report had the UAO comment, indicating unresolved taxa. The imbalance in discordant pairs (13 vs 1) was significant (McNemar’s exact p = 0.0018), supporting higher PCR sensitivity for GNRs when the smear indicated the presence of these rods. Focusing on the 13 PCR-positive/culture-negative cases, PCR identified 19 Gram-negative organisms not recovered by culture, including *Enterobacter cloacae*, *Klebsiella pneumoniae*, *Proteus mirabilis*, *Morganella morganii*, *Escherichia coli*, and *Stenotrophomonas maltophilia*. Organism-specific Ct values had a mean of 23.1, a median of 23.7, and a range of 16.5–31.3; 16/19 (84.2%) were Ct ≤ 28, and only one (*Stenotrophomonas maltophilia*, Ct 31.3) exceeded Ct 30.

**Figure 4.**
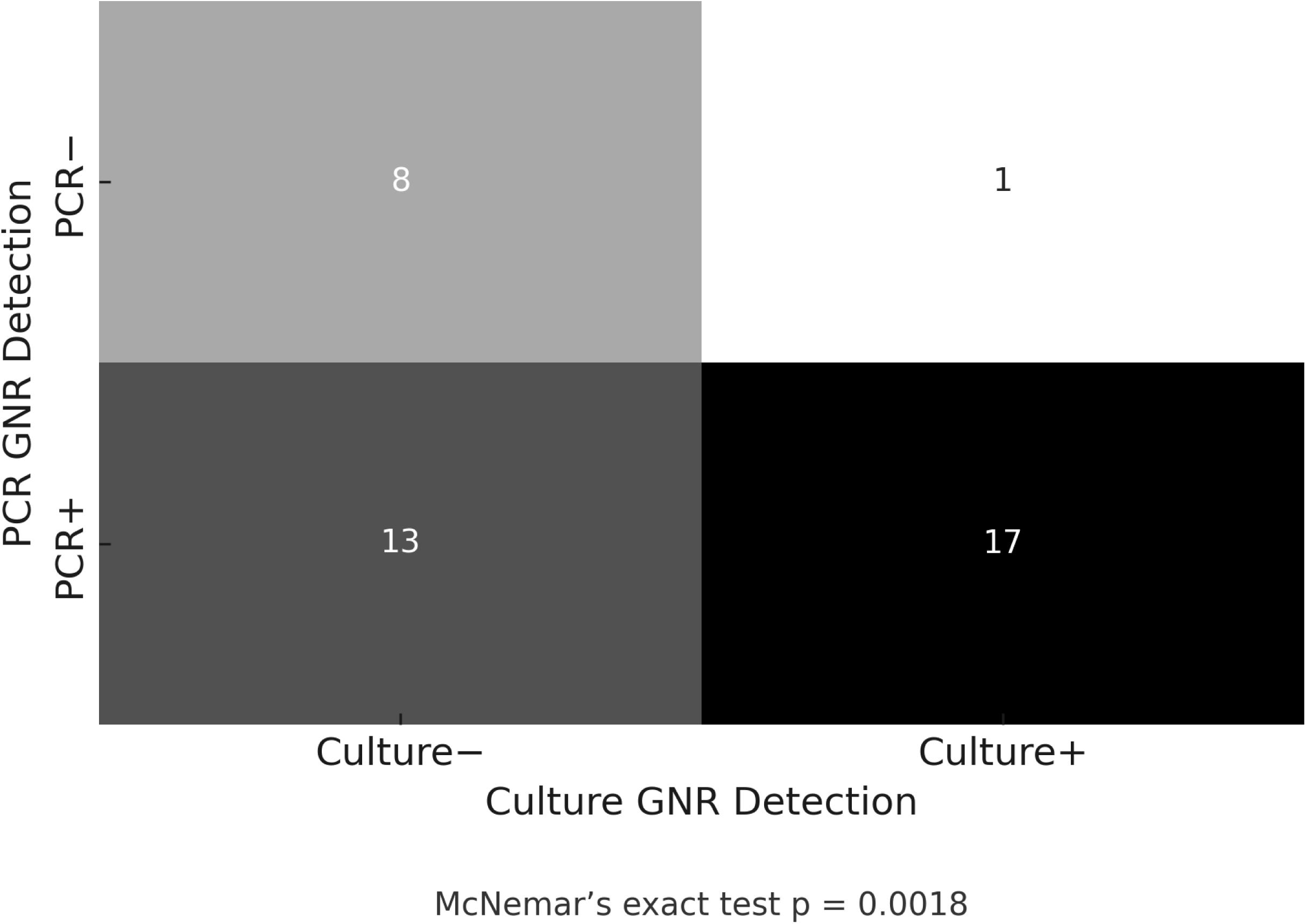
Heatmap summarizing concordance between PCR and culture for Gram-negative rod (GNR) detection among 39 cases with Gram stain–positive findings. PCR and culture both detected GNRs in 17 cases; 13 cases were PCR-positive but culture-negative, and only one was culture-positive but PCR-negative. The McNemar exact test p value = 0.0018, indicating the directional discordance between PCR (n = 13) and Culture (n = 1) is statistically significant.

To better evaluate the biological plausibility of organisms detected by PCR, in the absence of culture confirmation, we examined the co-detection of antibiotic resistance (ABR) genes and their related pathogens, using Ct value correlations as indirect indicators. This finding is illustrated in **Figure 5**, where each point represents an organism–gene pair (e.g., *DHA* with *Morganella morganii* (*18*), chromosomal *ampC* with *Enterobacter cloacae* (*19*), and *OXA*-group carbapenemases with *Acinetobacter baumannii* (*20*)). Tight clustering at low Ct values for pathogen-ABR pair datapoints suggests co-amplification from the same DNA source; dispersion increases at higher Ct (lower nucleic-acid abundance) and is consistent with low-template stochasticity.

**Figure 5.**
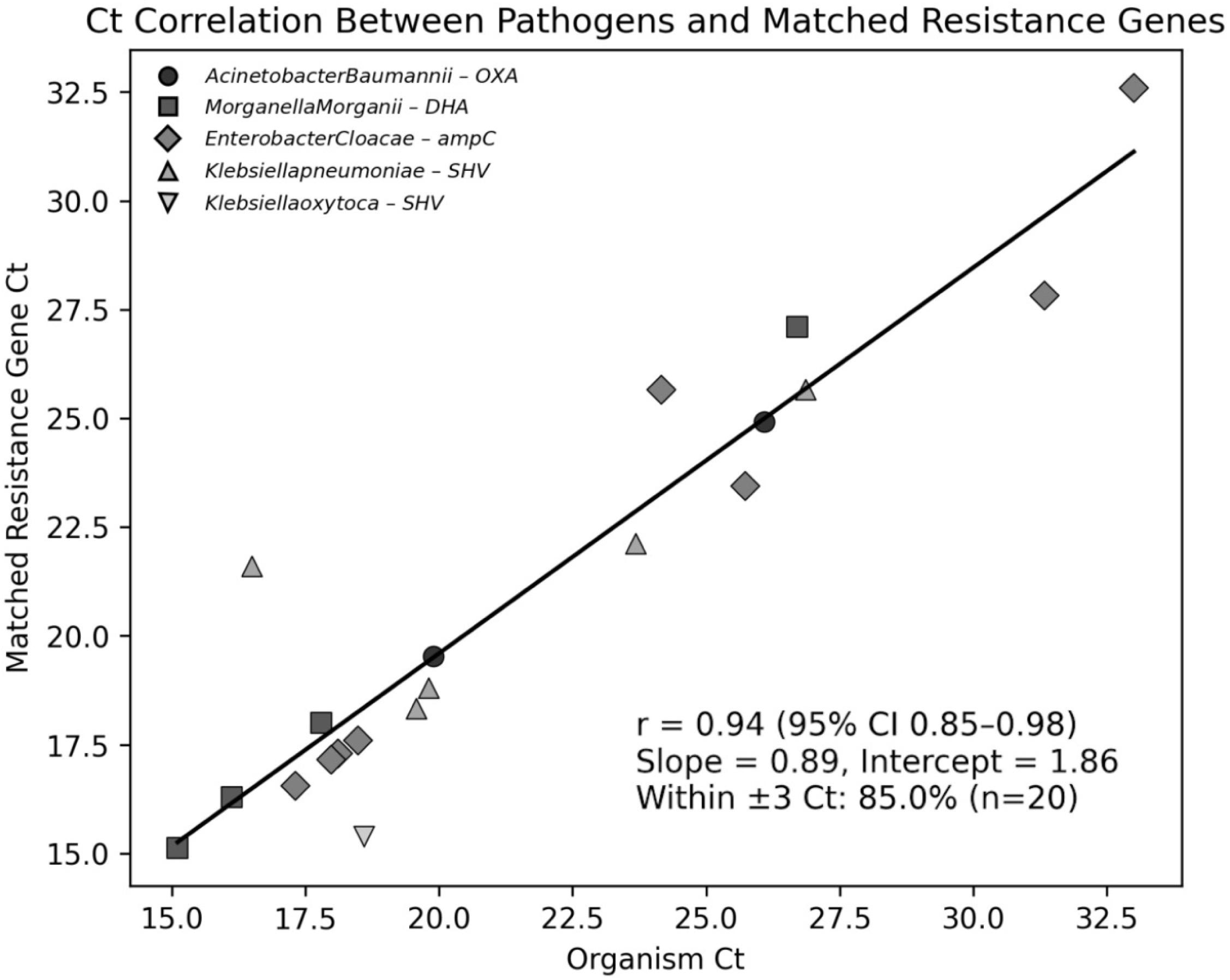
Ct correlation between PCR-detected pathogens and their matched resistance genes. This Scatter plot shows the relationship between the PCR Ct values of the detected Gram-negative organisms and their corresponding antibiotic resistance genes, with each point representing a biologically linked microorganism–ABR gene pair. Notable examples include *OXA* with *Acinetobacter baumannii*, *DHA* with *Morganella morganii*, *SHV* with *Klebsiella* species, and chromosomal *ampC* with *Enterobacter cloacae*. strong linear correlation was observed (Pearson r = 0.94; 95% CI, 0.85–0.98), with 85% of pairs falling within ±3 Ct. The regression slope of 0.89 suggests that pathogen and corresponding resistance gene abundances trend closely, supporting biological co-detection and reducing the likelihood of spurious amplification

### Clustering of "false positives" with infections

Within the conventional classification, comprising culture-confirmed positives including TP, TP- UAO, and Partial-TP cases (TP-all; n = 58), PCR-positive/culture-negative cases labeled as false positives (FP; n = 9), and true negatives, excluding 3 cases that were positive for organisms that were not covered by the PCR panel (TN; n = 22), we evaluated bacterial burden using *16S rRNA* Ct values (Ct_16_S). The median *16S rRNA* Ct was 15.44 for all TPs, 15.97 for FPs, and 23.60 for TNs. FP cases were not significantly different from all TP group (*p = 0.562*), indicating that PCR-positive/culture-negative results clustered with true infections in terms of bacterial load. In contrast, FP cases had significantly lower Ct values than TN (*p = 0.016*), reflecting a notably higher microbial burden than non-infected samples. Interestingly, the distribution of *<u>16S rRNA</u>* <u>Ct</u> values among false-positive (FP) cases appeared bimodal **(Figure 6**). One peak aligned with the high-burden region (Ct ≈ 15–18) observed in true positive groups, while a second smaller peak emerged around Ct ≈ 24–27, approaching the distribution of true negatives. This bimodal distribution suggests that cases labeled as FP using conventional classification are not microbiologically uniform. Such patterns support the interpretation that a subset of these so-called false positives may reflect true infections that were not captured by the culture method.

**Figure 6.**
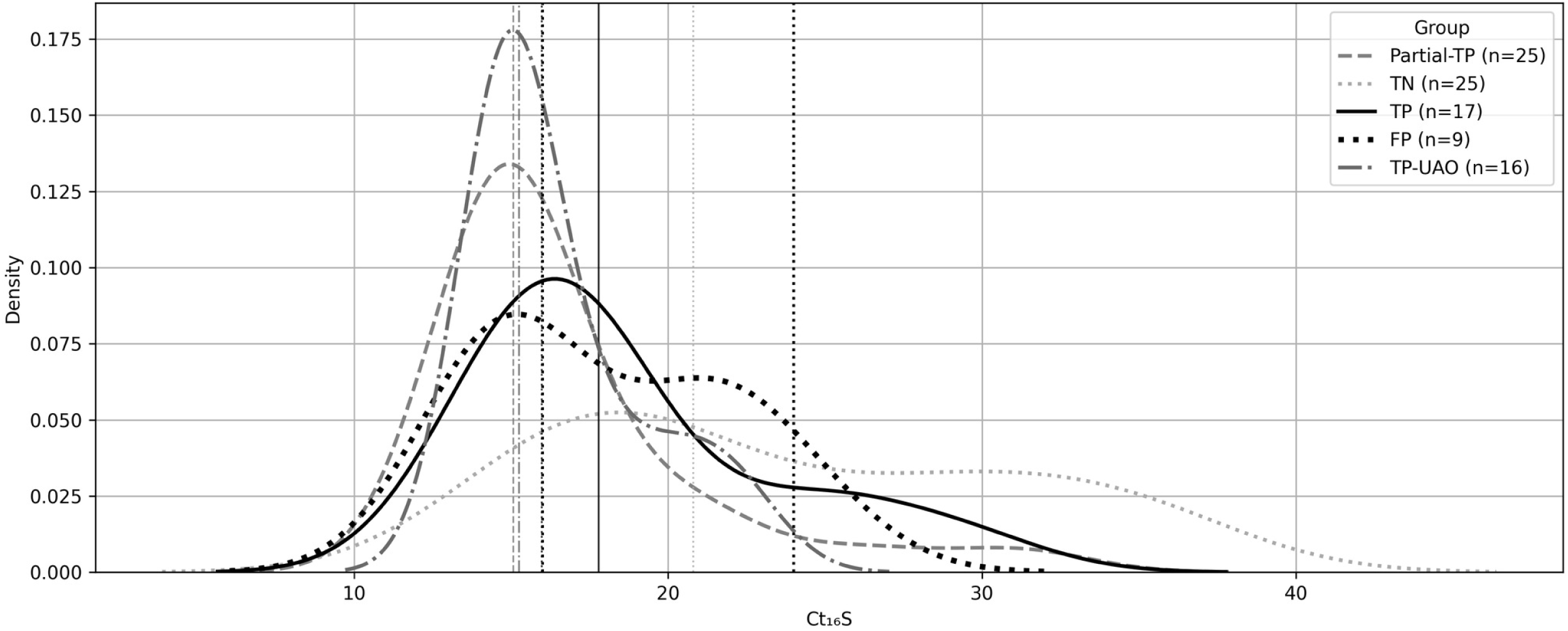
*16S rRNA* Ct values Kernel Density Distribution by Diagnostic Group. Distribution of pan-bacterial *16S rRNA* Ct values (Ct_16_S) across diagnostic categories under the conventional classification model. Lower Ct reflects higher bacterial burden. True positives (TP), Partial-TP, and TP-UAO cases show tightly clustered peaks at low Ct values (13–18), consistent with high biomass infections. False positive (FP) cases display a bimodal distribution: a dominant peak overlapping with TP groups (Ct ∼15–22) and a minor shoulder extending toward the TN range. True negatives (TN) show a broader, right-shifted distribution centered around higher Ct values (∼28–32), consistent with low microbial burden or background DNA. Line styles distinguish groups; vertical dashed lines mark estimated peaks for FP-high and FP-intermediate subclusters.

### Evaluation of infection status at the case-level using logistic regression modeling

To further assess PCR’s diagnostic validity, especially for cases classified as false positives (PCR+/Culture–), under the conventional classification model, we built multiple case-level logistic regression models integrating pan-bacterial burden *16S rRNA Ct* (Ct_16_S) and Gram smear morphology. Across all 93 included cases, Pan-bacterial *Ct_16_S* model alone predicted infection (β = –0.170; p = 3.8 × 10⁻⁵), with an odds ratio of 1.19 per one-cycle decrease (95% CI 1.09– 1.32) and AUC = 0.75, consistent with a biologically plausible link between higher bacterial burden and infection (Figure S3). We then restricted evaluation to an unambiguous 80-case subset (i.e., TP = 58, TN = 22; three TNs excluded for off-panel culture organisms). In this curated set, a Ct_16_S-only model achieved AUC = 0.847 (Figure 7). Adding morphology as binary indicators showed that GNR presence provided the most substantial incremental value: Ct_16_S + GNR reached AUC = 0.87, outperforming Ct_16_S + GPC (AUC = 0.83) and Ct_16_S + (GPC & GNR) (AUC = 0.845). Notably, Ct_16_S remained independently predictive across models (p < 0.001). In contrast, the presence or absence of individual Gram stain morphology was not significant (**Supplementary Table S9**), indicating that morphology improves ranking rather than adding orthogonal signal, which accords with the clinical tendency of GNRs to represent pathogens while some GPCs and GPRs reflect skin flora. Because binary flags only capture presence/absence, we next used semi-quantitative morphology (0–3 abundance scores for GPC and GNR), instead. The Ct_16_S + “Gram_Score_Sum” model (additive GPC+GNR index) performed best among scoring strategies with AUC = 0.862, exceeding “Gram_Score_Max” and “Gram_Score_GN_Priority” (both AUC = 0.842) and closely approaching the best binary model. We selected Ct_16_S + “Gram_Score_Sum” for probability estimation because it (i) integrates both morphologies into a single index; (ii) achieves near-maximal discrimination; and (iii) calibrated well. On out-of-fold (5-fold) evaluation, the AUC was 0.855 (raw 0.854), and calibration was close to ideal (intercept −0.053, slope 1.051). Applying the trained, calibrated Ct_16_S + “Gram_Score_Sum” model to the nine conventional FPs (Supplementary Table S10) produced calibrated probabilities spanning 0.48–0.90; 4 cases out of 9 had predicted probabilities ≥0.80, 7/9 were ≥0.70, and the two lowest were 0.59 and 0.48. Seven of nine FP cases had low *16S rRNA* (Ct ≤ 22), indicating high pan-bacterial burden, with Gram morphologies concordant with detected taxa (e.g., *Staphylococcus lugdunensis*, *Enterococcus faecalis*, *Citrobacter koseri*, *Staphylococcus aureus*). Cases Nos. 44, 30, and 33 are examples that the reference laboratory’s aerobic culture categorized growth as "skin flora only" (no recovery of *S. aureus*, *S. agalactiae*, or *P. aeruginosa*) despite concordant molecular and Gram findings. Case 44 (conventional FP; predicted infection probability >80%), PCR detected *Aerococcus urinae* (Ct 27.2), *Finegoldia magna* (Ct 23.8), and *Peptoniphilus* spp. (Ct 24.3), consistent with high bacterial load. Gram stain showed moderate Gram-positive cocci. Histopathological examination from the same site indicated acute osteomyelitis characterized by necrotic bone tissue and neutrophilic infiltrates. Case 30 (predicted infection probability > 85%); PCR detected *Staphylococcus lugdunensis* (Ct 16.1), *F. magna* (Ct 25.1), and CoNS (Ct 15.9), all at high abundance. Gram stain showed abundant Gram-positive cocci with no epithelial cells (high-quality sampling). Case 33 (predicted infection probability > 85%); PCR detected *S. lugdunensis* (Ct 21.9), *F. magna* (Ct 21.2), and CoNS (Ct 17.7). Gram smear demonstrated heavy Gram-positive cocci in pairs and clusters. *Staphylococcus lugdunensis*, although coagulase-negative, is a recognized pathogen in soft-tissue and bone infections and is often overlooked or misidentified as benign CoNS or skin flora (21).

**Figure 7.**
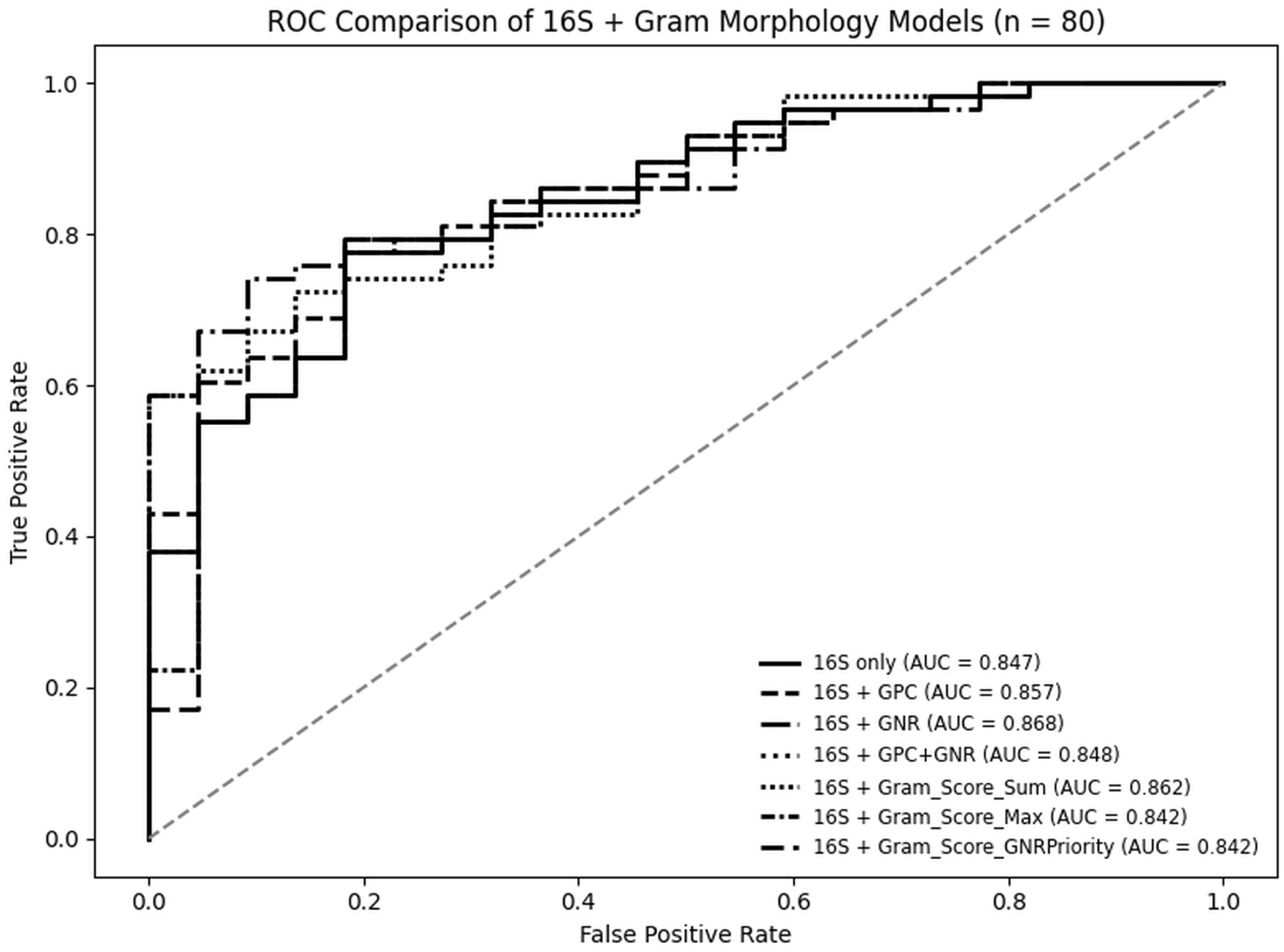
Receiver operating characteristic (ROC) curves for logistic regression models integrating *16S rRNA* Ct values and Gram stain morphology in wound infection diagnostics (n = 80). Models evaluated included *16S rRNA-*only, *16S rRNA* plus individual Gram morphology flags (GPC, GNR), and three scoring-based strategies: “Gram_Score_Sum”, “Gram_Score_Max”, and “Gram_Score_GNR Priority”. Among models incorporating both GPC and GNR Gram morphologies, the “*16S rRNA* + Gram_Score_Sum” model demonstrated the highest discrimination (AUC = 0.862) All models were trained using 80 curated specimens (TP and TN) and plotted in black and white for clarity.

### Evaluating the infection likelihood of PCR-detected organisms using biologically anchored models

We developed a logistic regression model to estimate the probability that a PCR-detected organism represents a true infection, integrating three biologically interpretable features: (i) organism-specific Ct (target abundance), (ii) pan-bacterial load (Ct_16_S), and (iii) Gram morphology. Logistic regression offers a transparent and interpretable framework that utilizes continuous biological variables, such as pathogen-specific burden and morphological context, and is widely employed in clinical prediction modeling (22). To preserve biological fidelity, only the morphology matching the organism’s expected Gram class was used for that organism (e.g., E. coli used the GNR score; the GPC score was set to 0). The primary aim of this Ct- and Gram morphology-integrated modeling approach was not to reclassify detections based on fixed thresholds, but to provide a statistically grounded, unbiased framework for evaluating the credibility of PCR-only results. Unlike rule-based contextual models that incorporate heuristic or subjective assumptions, this model was developed entirely from our culture-confirmed digitized data, without prior exposure to PCR-only detections.

Verified organism-level true positives (culture-matched organisms) and true negatives were used to train an L2-regularized logistic regression model. Platt scaling was applied using 5-fold cross-validation to calibrate the model’s probability outputs to obtain unbiased probability estimates for assessment (23). PCR-only detections were not included in the training data to minimize overfitting. Internal validation demonstrated excellent model performance, with an area under the receiver operating characteristic (ROC) curve (AUC) of 1.00, an overall accuracy of 99.9%, and an F1 score of 0.98 for the positive class. A high AUC value and F1 score were anticipated because the training set contained only verified biological input data. The model’s calibration curve demonstrated strong alignment between predicted probabilities and observed outcomes, confirming the accuracy of the probability estimates. To assess the model’s specificity in uninfected cases, we conducted simulations using 22 cases classified as true negatives under the standard diagnostic framework. For each case, we introduced a simulated high–Ct detection (Ct = 35) representing either Gram-positive cocci (GPC) or Gram-negative rods (GNR), while preserving the actual Gram morphology scores for the specimen. Under these conditions, the highest predicted infection probabilities observed were 0.541 for GPC and 0.221 for GNR, reflecting the model’s baseline noise level of low-burden signals in morphologically relevant but non-infectious contexts.

The trained, calibrated model was applied to all PCR-detected organisms (**Supplementary Table S11**). Among the PCR-only detections, 73.8% (71/103) had probability scores greater than 0.75. These findings suggest that most PCR-only organisms exhibit biological features typical of confirmed infections. Interestingly, among the 33 GNR+/Culture- cases, only 4/33 (12.1%) had a probability score below 0.75; their PCR Ct values ranged from 26.8 to 33.0. Additionally, 64% of these cases had probability scores higher than 0.90 (Table 6). Organisms such as *Morganella morganii*, *Proteus mirabilis*, *Citrobacter freundii*, and *Klebsiella oxytoca* ranked among the top predictions, with model probabilities approaching or equal to 1.00. These detections were characterized by low organism-specific Ct values (i.e., high burden), low *16S rRNA* Ct values (high pan-bacterial burden), and Gram-negative rod morphology, mirroring the biological signatures of confirmed pathogens, despite negative culture reports.

*Enterococcus faecalis* was also another organism of interest. Although it was not recovered by culture in the samples included in this study, it was detected with low Ct values in multiple cases and often scored above the 0.8 high-confidence threshold. As previously mentioned, *Enterococcus faecalis* was detected in 36 cases out of 107 specimens. However, in 93 cases included in the modeling studies, a total of 33 cases were positive for *Enterococcus faecalis* (n = 29) and *E. faecium* (n = 4). Predictive modeling provided strong probabilistic evidence supporting true infection in these cases. In the subset of cases evaluated for infection probability, calibrated scores ranged from 0.285 to 0.966, with a median probability of 0.639. As shown in Table 4, 81.8% (27/33) of these cases had probabilities ≥ 0.80, indicating high biological likelihood. These cases were characterized by Ct values < 28, Gram-positive cocci morphology on stain, and co-detection of resistance genes such as *van*, *ermA/ermB*, or *tet(M)*. The presence of vancomycin resistance genes was confirmed in 4 cases, further strengthening the clinical relevance of these detections. All *Vancomycin-resistant Enterococci* had infection probabilities of higher than 0.66. These results show that these *Enterococci* detections are not necessarily low-level noise or background colonizers but represent likely true positives with quantifiable biological support, likely missed due to culture’s analytical limitations, such as subjectivity or organism prioritization.

In case 7, a specimen obtained from a non–pressure ulcer on the right second digit showed the presence of white blood cells on Gram stain, despite a negative culture result. PCR detected *Enterococcus faecalis* with a Ct of 22.6 and a total bacterial burden (*16S rRNA* Ct) of 22.1, corresponding to a predicted probability of true positivity of 0.807 (Table 4). A macrolide resistance gene (*ermA/ermB*) was also identified with a Ct of 21.6. These findings enhance the clinical relevance of this PCR-only detection and suggest that the negative culture result may not reflect the true status of the submitted specimen. As summarized in Table 5, case no. 93 illustrates how molecular diagnostics can uncover a more complex and clinically meaningful pathogen profile than traditional culture alone. While culture identified *Pseudomonas* aeruginosa (moderate growth, susceptible to ciprofloxacin and levofloxacin), *Staphylococcus aureus* (light growth), and *Streptococcus agalactiae* (heavy growth), it also noted the presence of "additional organisms of unknown significance" that were not further characterized, suggesting an incomplete picture of the microbial profile.

**Table 5.** Example Organism Detections and Calibrated Infection Probabilities.

| Case | Organism | Ct | Ct_16S | GPC | GPR | GNR | Culture Positive? | Calibrated_Prob |
| --- | --- | --- | --- | --- | --- | --- | --- | --- |
| 1 | *E. cloacae* | 33.0 | 14.8 | Low | Absent | High | False | 0.636 |
| 1 | *E. faecium* | 31.3 | 14.8 | Low | Absent | High | False | 0.717 |
| 1 | *E. coli* | 16.8 | 14.8 | Low | Absent | High | True | 0.98 |
| 7 | *E. faecalis* | 22.6 | 22.07 | Absent | Absent | Absent | False | 0.807 |
| 93 | *E. coli* | 24.6 | 14.96 | Low | Absent | High | False | 0.903 |
| 93 | *S. aureus* | 30.1 | 14.96 | Low | Absent | High | True | 0.759 |
| 93 | *P. aeruginosa* | 26.3 | 14.96 | Low | Absent | High | True | 0.867 |
| 93 | *S. agalactiae* | 28.9 | 14.96 | Low | Absent | High | True | 0.8 |
| 93 | *E. faecalis* | 27.2 | 14.96 | Low | Absent | High | False | 0.85 |
| 93 | *K. oxytoca* | 22.4 | 14.96 | Low | Absent | High | False | 0.936 |
| 93 | *K. pneumoniae* | 19.8 | 14.96 | Low | Absent | High | False | 0.962 |
| 93 | *C. freundii* | 17.7 | 14.96 | Low | Absent | High | False | 0.975 |

**Table 6.** PCR-only Gram-negative Rods (GNRs) and Model-Inferred Infection Probabilities.

| No | Case no | Organism | Ct | Ct_16S | GNR | Culture Positive | Calibrated_Prob |
| --- | --- | --- | --- | --- | --- | --- | --- |
| 1 | 46 | *M. morganii* | 15.1 | 14.4 | High | FALSE | 0.987 |
| 2 | 55 | *K. pneumoniae* | 16.5 | 13.9 | High | FALSE | 0.984 |
| 3 | 95 | *E. cloacae* | 17.3 | 14.62 | High | FALSE | 0.978 |
| 4 | 93 | *C. freundii* | 17.7 | 14.96 | High | FALSE | 0.975 |
| 5 | 89 | *E. cloacae* | 18.0 | 14.78 | High | FALSE | 0.974 |
| 6 | 58 | *M. morganii* | 17.8 | 15.15 | High | FALSE | 0.974 |
| 7 | 104 | *K. oxytoca* | 18.7 | 14.25 | High | FALSE | 0.973 |
| 8 | 25 | *E. cloacae* | 18.1 | 14.98 | Low | FALSE | 0.973 |
| 9 | 47 | *P. mirabilis* | 18.0 | 15.16 | High | FALSE | 0.972 |
| 10 | 29 | *C. koseri* | 19.8 | 13.99 | Low | FALSE | 0.967 |
| 11 | 46 | *S. marcescens* | 19.9 | 14.4 | High | FALSE | 0.964 |
| 12 | 41 | *K. pneumoniae* | 19.6 | 15.07 | High | FALSE | 0.962 |
| 13 | 93 | *K. pneumoniae* | 19.8 | 14.96 | High | FALSE | 0.962 |
| 14 | 58 | *P. mirabilis* | 20.0 | 15.15 | High | FALSE | 0.959 |
| 15 | 19 | *P. mirabilis* | 22.9 | 13.68 | Low | FALSE | 0.942 |
| 16 | 93 | *K. oxytoca* | 22.4 | 14.96 | High | FALSE | 0.936 |
| 17 | 45 | *K. pneumoniae* | 23.7 | 15.34 | High | FALSE | 0.913 |
| 18 | 52 | *E. cloacae* | 24.2 | 14.75 | High | FALSE | 0.912 |
| 19 | 58 | *E. coli* | 24.0 | 15.15 | High | FALSE | 0.91 |
| 20 | 104 | *S. maltophilia* | 25.0 | 14.25 | High | FALSE | 0.906 |
| 21 | 93 | *E. coli* | 24.6 | 14.96 | High | FALSE | 0.903 |
| 22 | 19 | *M. morganii* | 26.8 | 13.68 | Low | FALSE | 0.879 |
| 23 | 19 | *P. vulgaris* | 27.2 | 13.68 | Low | FALSE | 0.87 |
| 24 | 89 | *P. aeruginosa* | 26.5 | 14.78 | High | FALSE | 0.866 |
| 25 | 52 | *K. pneumoniae* | 26.9 | 14.75 | High | FALSE | 0.857 |
| 26 | 46 | *C. koseri* | 27.8 | 14.4 | High | FALSE | 0.841 |
| 27 | 29 | *K. oxytoca* | 29.5 | 13.99 | Low | FALSE | 0.8 |
| 28 | 56 | *S. maltophilia* | 26.5 | 18.4 | High | FALSE | 0.779 |
| 29 | 15 | *P. mirabilis* | 30.3 | 14.13 | High | FALSE | 0.769 |
| 30 | 57 | *K. oxytoca* | 29.8 | 17.08 | High | FALSE | 0.694 |
| 31 | 48 | *S. maltophilia* | 31.3 | 15.75 | High | FALSE | 0.677 |
| 32 | 98 | *K. oxytoca* | 26.8 | 21.18 | Low | FALSE | 0.676 |
| 33 | 1 | *E. cloacae* | 33.0 | 14.8 | High | FALSE | 0.636 |

In contrast, the PCR test not only confirmed the organisms identified by culture but also detected several additional pathogens with low Ct values, including *Citrobacter freundii* (Ct 17.7), *Morganella morganii*, *Klebsiella pneumoniae* (Ct 19.8), *Klebsiella oxytoca* (Ct 22.4), *Escherichia coli* (Ct 24.6), and *Enterococcus faecalis* (Ct 27.2). Interestingly, the calibrated detection probabilities for *M. morganii*, *K. pneumoniae*, *K. oxytoca*, and *E. coli* exceed those of organisms also confirmed by culture, likely because their Ct values are lower than those of the other organisms. Importantly, multiple antimicrobial resistance (ABR) genes were also identified: *BIL/LAT/CMY* (Ct 17.8), *SHV* (Ct 18.9), and the *qnr* gene group (Ct 23.6). Although PCR cannot definitively assign these resistance markers to specific organisms, the proximity of Ct values combined with known organism-ABR gene relationships suggests that *BIL/LAT/CMY* is most likely linked to *C. freundii*, *SHV* to *K. pneumoniae*, and the *qnr* group to *E. coli*. Notably, the Ct value for *Pseudomonas aeruginosa* (26.3) was higher than those of other Gram-negative organisms, indicating that it was not the primary source of infection and that the *qnr* gene probably originated from *E. coli*. Overlooking this resistance could lead to a targeted treatment based solely on culture AST on *Pseudomonas aeruginosa*, which may be suboptimal or ineffective.

Another illustrative example is case No. 1 (**Table 5**), where *Enterococcus faecium* was detected by PCR with a Ct value of 31.3, and a moderate predicted infection probability of 0.717. However, the concurrent detection of the vancomycin resistance gene (*vanA/vanB/vanC*) with a Ct of 31.25 offers additional support for the clinical relevance of this finding, despite its low abundance. In this case, the culture report indicated heavy growth of *E. coli*, which aligns perfectly with the PCR result (Ct 16.8). Interestingly, in two cases (10, 100), *Staphylococcus aureus* was detected by both PCR and culture, yet the model assigned low infection probabilities (0.506 and 0.178, respectively). While both were technically true positives, they lacked the strong biological signature typically associated with high-confidence calls. *Staphylococcus aureus* Ct values were 28.9 and 31.3, respectively.

In contrast, all detections of *Pseudomonas aeruginosa* in this study fell within the high-probability range, reflecting consistently low Ct values, strong 16S burden, and presence of Gram-negative rods in Gram stain, highlighting the model’s ability to distinguish strong from weak biological signals in an unbiased manner. It should be noted that this model was not designed as a diagnostic classifier, but rather as a framework to objectively quantify the biological plausibility of each detection, using continuous variables such as Ct, total bacterial load, and Gram morphology score. In this context, lower infection probabilities indicate less compelling biological support, but not necessarily diagnostic irrelevance, and help prioritize the most confidently detected PCR-only organisms.

Together, these examples highlight the clinical significance of this PCR method in elucidating the profile of polymicrobial infections that may be overlooked during colony isolation or culture prioritization strategies. Additionally, this PCR Ct-, Gram morphology-integrated logistic regression model offers a robust and unbiased framework for assessing PCR-only detections. These findings suggest that most PCR-only detections in our cohort appeared to represent true infections rather than laboratory artifacts.

### Probability-adjusted symmetric confusion matrices support the validity of organism-level diagnostic accuracy in the PCR Panel

To further assess organism-specific diagnostic performance, we conducted a direct comparison of organism-level sensitivity and specificity for organisms detected by PCR using a symmetric reclassification model and the traditional culture-referenced framework. The symmetric confusion matrices at the organism level, in which each test was evaluated using the other as a reference, avoid the biases inherent in gold-standard-based metrics by penalizing both PCR and culture equally for discordant results. However, one limitation of symmetric matrices is that they treat all disagreements equally, without considering biological context (24), such as organism burden and/or semi-quantitative co-presence of relevant Gram morphology. Therefore, to improve the model in an unbiased manner, we integrated the organism-level probability-based diagnostic model discussed in the previous subsection to estimate the likelihood that a given PCR-detected organism represents a true infection. As mentioned earlier, this biologically informed probability score was used to reclassify high-confidence PCR-only detections (≥ 0.80) as true positives for PCR and false negatives for culture within the symmetric framework. This hybrid approach preserves the fairness of symmetric evaluation while accounting for biological plausibility and the burden on microorganisms, thereby providing a more nuanced and evidence-driven estimate of diagnostic performance.

We focused on five clinically significant pathogens that have higher detection prevalence in culture and/or PCR data: *Staphylococcus aureus*, *Pseudomonas aeruginosa*, *Streptococcus agalactiae*, *Escherichia coli*, and *Bacteroides fragilis*. Under this hybrid symmetric classification approach, PCR sensitivity for these five organisms ranged from 87% to 100%, and the specificity range was 91% to 100**% (Supplementary Table S12**).

For example, *Staphylococcus aureus* showed a sensitivity of 97.3% and a specificity of 94.1% under this symmetric reclassification model, compared to 97.2% and 91.4%, respectively, when using culture results as the gold standard. *Pseudomonas aeruginosa* showed sensitivity of 87.5% and specificity of 100% with symmetric modeling, compared to 86.7% and 98.2% using culture as the reference. *Streptococcus agalactiae* achieved perfect sensitivity (100%) and a specificity of 93.8% with reclassification; specificity declined slightly to 92.4% under the gold-standard model due to five PCR-only detections not recovered by culture. These trends were consistent across other high-prevalence organisms, such as *E. coli* and *B. fragilis*, for which PCR achieved 100% sensitivity and 95–100% specificity under both comparison approaches. These findings are aligned with earlier analyses in this study. In a targeted review of 28 wound specimens in which culture definitively ruled out *Staphylococcus aureus*, *Pseudomonas aeruginosa*, and *Streptococcus agalactiae*, PCR showed 100% specificity for the latter two organisms and a single detection of *Staphylococcus aureus*, yielding an overall specificity of 96.4%. In the whole 77-case subset group, the specificity for these three pathogens remained consistently high (96.9%), with statistical support from both binomial testing and Bayesian modeling (posterior mean: 96.4%, 95% CrI: 93.5%–98.6%). Logistic regression modeling further confirmed high infection probability for the majority of these detections, with 81.8% of *Enterococcus faecalis* and *E. faecium* entries scoring ≥ 0.80. With this hybrid symmetric modeling, *Enterococcus faecalis* showed a sensitivity of 100% and specificity of 91%, compared to 0% and 100% using culture as the reference.

### Tiered evaluation of PCR and culture performance using predicted probabilities and ABRs

To comprehensively evaluate the diagnostic performances of culture and PCR within the context of our initial conventional classification, while considering organism-level concordance, we implemented a tiered reclassification framework comparing PCR and culture using three progressively refined models. Each model defined true positive (TP), false positive (FP), false negative (FN), and true negative (TN) designations at the case level, based on the concordance or discordance between PCR and culture, and guided by Organism Ct- and Gram-morphology calibrated likelihood and co-detection of relevant ABR genes. We began by evaluating case-level diagnostic performance using the penalized PCR classification as a baseline, where culture served as the comparator. In this classification strategy, the detection of any organism not reported by culture led to classification as a false positive for PCR, even in cases where biological plausibility may have supported the detection. This resulted in a total of 34 true positives (TP), 31 false positives (FP), three false negatives (FN), and 25 true negatives (TN), corresponding to a sensitivity of 92%, a specificity of 45%, a positive predictive value (PPV) of 52%, and an overall accuracy of 63% (**Table 7 and Supplementary Table S13**); It should be noted that, in this specific framework, cases whose culture reports contained partially informative findings due to “unidentified additional organisms,” but included organisms matching PCR detections (TP UAO), were classified as acceptable true positives (just for penalized PCR framework) to ensure that the overall case count for all models remains consistent (n = 93). To account for biologically and or clinically plausible organisms detected by PCR but not recovered by culture, we next applied a Likelihood-Adjusted (LA) classification framework. In this model, PCR-only organisms were retained as true positives if they met predefined infection probability thresholds (≥ 0.8 for Gram-positive cocci; ≥ 0.5 for Gram-negative rods), derived from the organism’s Ct and morphology-informed logistic regression model discussed in the previous section. These thresholds were determined by applying a 50% relative increase over the estimated noise levels (0.54 for GPCs, 0.21 for GNRs). PCR organisms meeting these criteria, especially when supported by low Ct values and high Gram morphology scores, were considered biologically credible infections. This adjustment reduced the number of false positives from 31 to 11 while increasing true positives to 54. Sensitivity remained 95%, and specificity improved to 69%, with overall accuracy rising to 85%. Subsequently, in the LA-Culture model, 25 cases retained their true positive label, and likely false negative counts increased to 40, resulting in 100% specificity and PPV, but a sensitivity of 38% and overall accuracy of 57%. The ABR + Likelihood-Adjusted (PCR) model further refined this classification by incorporating antimicrobial resistance (ABR) gene detection as a tie-breaker for organisms with intermediate infection probability scores (defined as 0.60–0.80 for GPCs and 0.30–0.50 for GNRs). Under this framework, an additional 7 cases were reclassified as likely true positives, increasing the TP count to 61 and reducing false positives to 4. The resulting specificity increased to 86%, and accuracy reached 92%, with no change in sensitivity or NPV. Subsequently, in the ABR + LA-Culture model, TP cases with TP label were reduced to 23, and false negative counts increased to 45, resulting in 100% specificity and PPV, but a sensitivity of 34%, an NPV of 36% and overall accuracy of 52%. These results suggest that this culture-based method may underrepresent some of the known clinically significant pathogens, particularly in cases involving polymicrobial infection.

**Table 7.** Diagnostic Performance Metrics Across Unadjusted PCR (penalized PCR), Probability-Adjusted, and ABR-Informed & Probability-Adjusted Models (n = 93)

| Model | TP | FP | FN | TN | Sensitivity | Specificity | PPV | NPV | Accuracy |
| --- | --- | --- | --- | --- | --- | --- | --- | --- | --- |
| Penalized PCR (baseline) | 18 +16* | 31 | 3 | 25 | 0.92 | 0.45 | 0.52 | 0.89 | 0.63 |
| Likelihood-Adjusted (PCR) | 54 | 11 | 3 | 25 | 0.95 | 0.69 | 0.83 | 0.89 | 0.85 |
| ABR + Likelihood-Adjusted (PCR) | 61 | 4 | 3 | 25 | 0.95 | 0.86 | 0.94 | 0.89 | 0.92 |
| Likelihood-Adjusted (Culture) | 25 | 0 | 40 | 28 | 0.38 | 1.0 | 1.0 | 0.41 | 0.57 |
| ABR + Likelihood-Adjusted (Culture) | 23 | 0 | 45 | 25 | 0.34 | 1.0 | 1.0 | 0.36 | 0.52 |

\*TP-UAO cases were labeled as TP to include all 93 cases in the calculation; \* To ensure full transparency, we also report the expected performance shift if off-panel organisms—i.e., culture-only detections not included in the PCR target panel—are considered during evaluation. Under this expanded reference framework, one PCR false positive is reclassified as a false negative. This change results in a slight decrease in sensitivity (from 95.4% to 86.4%) but improves specificity (from 89.3% to 92.6%) for the ABR+ Likelihood-Adjusted PCR model. The culture-based performance metrics remain unchanged under this adjustment, as none of the culture-only organisms met model-based criteria for reclassification.

### Overall diagnostic performance evaluation using the Latent-class model fit

In the absence of a definitive reference standard, we also applied latent class analysis (LCA) to estimate the case-level diagnostic accuracy of PCR, culture, and Gram stain without assuming any single method to be perfectly accurate. LCA treats the true infection status as a latent (unobserved) binary variable and models the observed combination of test results at the case level across the 93 included specimens. All three tests, PCR, culture, and Gram stain, were available for every case, allowing us to estimate their performance characteristics jointly.

Out of the four candidate latent class models, only Models M1 and M3 converged. However, since M1 uses only two binary indicators (PCR and culture), the two-class LCA is not identifiable; multiple parameter sets result in essentially identical fits to the observed 2×2 table. Consistent with this, M1’s item-response estimates were sensitive to initialization and therefore are not interpreted further. Using the three-indicator latent class model (M3) and the panel-restricted definition, we estimated very high sensitivities for PCR (100%) and Gram stain (96.8%), with specificities of 90.6% and 32.9%, respectively; culture showed 91.0% sensitivity and 100% specificity. Model-based PPV and NPV are summarized in Table 8 (parametric bootstrap, seed = 2025). Results were similar under the all-organisms definition, with PCR sensitivity 95.6% and specificity 90.7%, culture sensitivity 92.0% and specificity 99.1%, and Gram stain sensitivity 97.0% with specificity 38.2%.

**Table 8.** Diagnostic accuracy estimates from latent class analysis (LCA: M3) under two definitions of culture positivity.

| SCENARIO | TEST | SENSITIVITY (%) | SPECIFICITY (%) | PPV (%) / NPV (%) |
| --- | --- | --- | --- | --- |
| PANEL-ONLY | PCR | 100 (100–100) | 90.6 (68–100) | PPV 95.8 / NPV 100.0 |
|  | Culture | 91.0 (81–100) | 100 (100–100) | PPV 100 / NPV 82.4 |
|  | Gram stain | 96.8 (92–100) | 32.9 (16–53) | PPV 77.0 / NPV 81.2 |
| ALL | PCR | 95.6 (89–100) | 90.7 (67–100) | PPV 96.7 / NPV 91.7 |
|  | Culture | 92.0 (82–100) | 99.1 (89–100) | PPV 98.8 / NPV 83.3 |
|  | Gram stain | 97.0 (92–100) | 38.2 (19–63) | PPV 79.9 / NPV 87.0 |

\*Note: Latent class model-based diagnostic performance (M3: PCR + Culture + Gram stain) under two analytic definitions of culture positivity. Values shown are sensitivity, specificity, positive predictive value (PPV), and negative predictive value (NPV), each with 95% confidence intervals derived from a 300-replicate parametric bootstrap (seed = 2025). The panel-only scenario considers only organisms targeted by the PCR assay, whereas the all-organisms scenario includes all recovered culture organisms, including off-panel organisms. Confidence intervals reaching 100% reflect boundary constraints of the parameter space.

## Discussion

This real-world comparison between an analytically validated, comprehensive PCR panel and a commercial reference laboratory’s culture demonstrates that PCR provides higher diagnostic sensitivity and wider pathogen detection for podiatric wound infections, especially in polymicrobial contexts, when both tests are performed in parallel on dual swabs from the same site. These findings are specific to these evaluated tests but align with prior work showing that molecular approaches reveal organisms that routine culture under-recovers in complex wounds. It should also be noted that multiple studies using different molecular techniques have documented the underdetection of anaerobes and fastidious bacteria, such as *Peptoniphilus* species, *Peptostreptococcus* species, *Finegoldia magna*, and *Actinotignum schaalii by* traditional culture (25–27). By excluding these organisms (defined as HTC organisms, Table S3), we primarily focused on the clinically significant pathogens that are expected to be detectable through both culture and PCR, if they are present in a collected specimen.

Within the culture-referenced conventional framework, PCR showed 98.3% sensitivity (58/59) and 73.5% specificity (25/34). It also detected 110 organisms not reported by culture. Discordance favored PCR at both the case (McNemar p = 0.021) and organism (p ≤ 1.2×10⁻⁶) levels, suggesting culture frequently missed or deprioritized clinically relevant taxa. Additional detections clustered in ∼30% of the 107 cases where culture reports noted ‘unidentified additional organisms’ (UAO) or were otherwise inconclusive, consistent with frequent polymicrobial infection in these podiatric specimens. This study presents multiple lines of biological evidence supporting the reliability of PCR-only additional detections.

First, 84% of PCR-only Gram-negative rods (GNRs) have organism-specific Cts ≤ 28. These detections were often correlated with the GNR morphology on Gram stain findings (**Supplementary Table S11**). Interestingly, among 39 specimens with a positive Gram stain smear for GNR morphology, the directional discordance in detecting GNR organisms favored PCR (McNemar p = 0.0018: statistically significant), indicating that PCR showed better correlation with Gram stain than culture even when the GNR level was abundant in the smear. This finding shows culture results do not align with the College of American Pathologists standard, MIC.11060, which emphasizes the role of Gram stain morphology in guiding culture workup and organism identification, especially when atypical or unexpected findings arise. Multiple studies have demonstrated that lower PCR cycle threshold (Ct) values, typically in the range of ≤ 28 to 30, are strongly associated with the presence of viable bacteria, as confirmed by culture, across diverse specimen types including stool, respiratory secretions, and wounds (28–31). The clinical impact of missed pathogens extends beyond underdiagnosis. In one illustrative case, culture identified Fluoroquinolone-sensitive Pseudomonas aeruginosa with moderate growth. Still, PCR detected five additional Gram-negative organisms with lower Ct values and co-detected three relevant ABR genes, including a fluoroquinolone resistance gene (*qnr*), likely originating from the PCR-only *E. coli*. This discrepancy suggests that therapy guided solely by culture, especially if the microbiology workup is not comprehensive, could be suboptimal, particularly in polymicrobial infections.

Second, microorganism–ABR gene pairs frequently demonstrated concordant Ct values (e.g., *ampC* with *Enterobacter cloacae*, *DHA* with *Morganella morganii*), indicating co-amplification from the same microbial DNA source.

Third, case- and organism-level modeling showed that higher bacterial burden (*16S rRNA Ct /* Organism Ct) and concordant Gram morphology increase the infection likelihood (Figure 7, Supplementary Table S10). The *16S rRNA* + Gram_Score_Sum model (additive GPC+GNR index) showed an AUC of 0.862; out-of-fold calibrated AUC was 0.855; calibration intercept was −0.053, with a slope of 1.051. When applied to the nine FP cases in the conventional classification framework, calibrated probabilities ranged from 0.48 to 0.90; four out of nine were ≥ 0.80, and seven out of nine were ≥ 0.70. For example, three "skin flora only" culture reports (Cases 44, 30, 33) exhibited high modeled probabilities alongside matching smear results with organisms detected by PCR. In Case 44, histopathologic evidence of acute osteomyelitis was observed (data coming from the histopathology report for the same site), indicating signals that are not consistent with background noise. These findings support previous research showing that combining quantitative molecular load with smear microscopy results can effectively distinguish true infections from colonization (32). We also employed latent class analysis (LCA) to evaluate case-level performance of the tests without treating any method as infallible. The chosen LCA model (which included PCR, culture, and Gram stain as independent indicators) yielded an estimated PCR sensitivity of ∼95.6% and specificity of ∼91%. These LCA-derived metrics support the conclusion that PCR offers analogous diagnostic performance compared to the culture test for detecting infected from non-infected specimens. Our work thus adds to the growing literature employing LCA as a helpful tool for objectively estimating the accuracy of molecular tests when traditional reference methods are imperfect (33–38). When the *16S rRNA* Ct + Gram_Score_Sum logistic regression model was applied to the nine conventional false positives, four yielded predicted infection probabilities ≥ 0.80. Under the conventional classification (Table 1: 58 TP, 9 FP, 1 FN, 25 TN), PCR demonstrated a sensitivity of 98.3%, specificity of 73.5%, PPV of 86.8%, and NPV of 96.3. Reclassifying the four high-probability cases as likely true positives would revise the counts to 62 TP, 5 FP, 1 FN, and 25 TN, improving specificity to 83.3% and PPV to 92.5%, while sensitivity and NPV remained high (98.4% and 96.2%). This adjusted estimate is directionally consistent with the LCA-derived results, further supporting that some PCR-only detections represent true infection rather than analytical or clinical false positives. However, it should be noted that this LCA approach only estimates each test’s ability to classify specimens as infected or non-infected and does not determine whether the complete and accurate pathogen profile was identified. Therefore, in the next step, we aimed to check the performance of PCR at the organism-level using a method-agnostic interpretive framework that integrates pathogen type (based on their Gram morphology), bacterial burden (organism Ct), and smear morphology scores, i.e., the Ct-informed, Gram morphology-aware organism-level model. This model indicated that most PCR-only detections show a biological signature very similar to the culture-matched PCR detections. To our knowledge, this is one of the first predictive models that simultaneously incorporates organism-specific Ct, total *16S rRNA* Ct, and Gram morphology scores to estimate organism-level infection likelihood. Previous studies have independently shown that low Ct values are associated with viable or clinically significant bacterial infections in wound, respiratory, and bloodstream samples (28, 30, 31), and others have shown improvement in pneumonia diagnostics by combining Gram stain with total bacterial burden using the *16S rRNA* Ct (32). When this model was applied to all PCR detections, 73.8% (71/103) of PCR-only detections yielded a calibrated likelihood of greater than 0.75. Among 33 GNR+/culture− detections, only 12.1% were below 0.75, and 64% were ≥ 0.90, driven by low organism-specific Cts, low *16S rRNA* Ct value, and higher GNR morphology smear scores, features mirroring culture-confirmed pathogens. *Enterococci* showed a clear pattern of under-recovery in the culture results for this cohort, which differs from Branch-Elliman et al., who reported that E. faecalis (149/859 = 17.3%) was the second most common organism in podiatric wound cultures, following S. aureus (339/859 = 38.8%) (39). However, the PCR test detected them in 35.5% of cases (*E. faecalis*: 29/93, and E. faecium: 4/93), with low Ct values, concordant GPC morphology, and often with codetection of *van/erm/tet* ABR genes; 81.8% (27/33) of Detected *Enterococci* had calibrated probabilities of ≥ 0.80. Median Ct for PCR-only *E. faecalis* calls overlapped with culture-confirmed *S. aureus’s Cts* (22.5 vs 19.2; Mann–Whitney p = 0.0128; KS p = 0.066), further arguing against low-level contamination or amplification of residual DNA from non-viable organisms.

To avoid penalizing PCR-only detections solely based on the assumption that culture is the gold standard, we also used symmetric confusion matrices at the organism level. Then we reclassified high-plausibility PCR-only detections (≥ 0.80) as PCR likely true positives and culture likely false negatives. Under this hybrid symmetric approach, PCR maintained high sensitivity and specificity (87–100% and 91–100%) for key pathogens, such as *S. aureus* (97.3%/94.1%), *P. aeruginosa* (87.5%/100%), *S. agalactiae* (100%/93.8%), concordant with organism-level culture-referenced performance analysis outlines in Table 2. These results are consistent with a PCR sensitivity of 100% and specificity of 90% for these three organisms in Melendez et al.’s study, where 39 specimens from 29 chronic wound infections were compared against culture (40). For *E. faecalis*, symmetric reclassification yielded 100% sensitivity and 91% specificity (vs 0%/100% using culture as reference), highlighting that this culture test likely missed detections rather than PCR, yielding false positive results. Targeted specificity checks (98.8% across 84 ruled-out events; 96.9% across 195 events in a 77-case subset) support the analytical precision for these organisms further. Collectively, these results indicate that issues like nonspecific amplification, background detection, or the detection of DNA from non-viable organisms are rare with PCR for these key pathogens, at least in this cohort of specimens. Therefore, most PCR-only detections are likely overlooked or missed by this culture method. In line with this statement, Cummings et al., reported that dominant pathogens with distinctive morphology, such as Staphylococcus aureus, often draw disproportionate attention during culture review, which can mask the presence of clinically important but less noticeable organisms or colonies (41).

To illustrate how case-level discordance between culture and the PCR method would look if we include all PCR-only additional detections, we compared three tiered frameworks based on the predicted probability of each PCR-only detection: (a) penalized PCR versus culture results, using culture as the reference, (b) a likelihood-adjusted classification that uses our Organism Ct and Gram score-informed regression model’s probabilities, and (c) an extended version of the likelihood model that incorporates evidence of antimicrobial resistance genes. Under the penalized PCR model (framework a), any additional organisms detected by PCR that were not detected in culture were counted against the specificity, yielding a low apparent specificity (∼45%) and an overall accuracy of only ∼63%. However, when we reclassified cases using the model-informed probabilities (framework b), PCR’s specificity improved markedly to ∼69% and accuracy to ∼85%, without loss of sensitivity. Adding the presence of relevant resistance genes as an extra layer of specificity (framework c) further increased PCR’s specificity to 86% and the overall diagnostic accuracy to 92%, while maintaining high sensitivity (∼95%).

In contrast, when we applied analogous reclassification logic to culture, and penalized culture for overlooking those likely TPs, culture’s specificity remained 100% but its sensitivity dropped to ∼38%. In other words, culture appeared "perfectly specific," while it missed a substantial portion of true organisms.

In summary, this study demonstrated that culture often underestimated pathogen diversity; among Gram-negative rods, organisms such as *Klebsiella* species, *Proteus mirabilis*, *Morganella morganii*, and *Acinetobacter baumannii*, and from Gram-positive cocci, *Enterococcus* species and *Staphylococcus lugdunensis* were often overlooked or missed by culture.

PCR showed higher concordance with GNRs on Gram smears (30/39) than culture (18/39); most of these PCR detections had Ct <28, consistent with moderate to high organism burden. This finding aligns with Gardner et al., who showed that culture underestimates microbial diversity in diabetic foot ulcers (42). In Hung et al.’s study of 558 patients, the presence of GNRs such as Escherichia coli, Proteus spp., and Pseudomonas aeruginosa was significantly linked to increased amputation risk in infected diabetic foot ulcers, emphasizing the importance of detecting all clinically relevant organisms in complex wound cases (43).

In our dataset, PCR delivered faster results (turnaround time favored PCR by >4 days (median ∼20 h for PCR vs ∼120 h for culture), and identified additional organisms supported by low Ct values, concordant Gram-stain morphology, and resistance-gene co-detection. Using a method-agnostic interpretive framework, we also demonstrated that many PCR-only detections exhibited biological signatures indistinguishable from culture-confirmed pathogens. Although culture remains central to diagnostic practice, it consistently underestimates the range of organisms present in complex specimens. Even organisms that are capable of growth may be missed because colony recognition is limited when culture plates contain many morphologies, dominant flora or fungi obscure minority populations, and technologists naturally focus on well-established pathogens (41). Studies in other specimen types confirm that sequencing methods recover a broader spectrum of cultivable organisms than culture alone (44). Our findings are consistent with these observations: PCR often detected pathogens that were not identified by standard culture methods, indicating that many presumed false positives in the penalized PCR model are indicative of clinically significant infections that culture methods failed to detect. The performance of culture techniques is limited by factors such as media selection, competitive growth, and subjective interpretation (41). In contrast, PCR offers a more inclusive, assumption-independent approach, enabling improved recognition of fastidious or resolving polymicrobial infections that traditional methods often overlook.

Taken together, PCR shows a more than 4-day faster turnaround time; LCA-estimated performance of approximately 95.6% sensitivity and approximately 91% specificity; ABR plus likelihood-adjusted performance of approximately 95% sensitivity, approximately 86% specificity, and 92% overall accuracy; and about 75% of PCR-only detections with high biological likelihood (Gram morphology and ABR support). These results suggest that, in this cohort, routine PCR detects clinically relevant pathogens that culture often under-detects in polymicrobial wounds and allows for earlier, more conclusive clinical decisions.

## Data Availability

All data produced in the present work are contained in the manuscript

## Acknowledgments

The authors thank the laboratory staff at BioExcel Diagnostics, LLC and Sagis Diagnostics, PLLC for their dedicated work in specimen processing, PCR testing, and quality control. We also acknowledge the contributions of the quality assurance team whose efforts supported the integrity and clarity of this study.

## Author Contributions

M.D. conceived the study. M.D. oversaw study design, data analysis, and manuscript preparation. H.N., S. D., M.G., contributed to data collection, analysis, interpretation, and critical manuscript review. H.L.M., M.G., and K.H.L. provided clinical oversight and critical manuscript review. M.L.S. contributed to the manuscript review. Authors read and approved the final manuscript.

## Conflict of Interest Statement

M.D. is the Chief Scientific Officer of BioExcel Diagnostics and a member of BioExcel Diagnostics, LLC. H.L.M., and K.H.L. are Medical Directors of Sagis PLLC and have indirect ownership in BioExcel Diagnostics, LLC.

## IRB Statement

This study was exempt from Institutional Review Board (IRB) oversight because it only involved retrospective analysis of de-identified laboratory reports as part of internal quality assurance efforts at BioExcel Diagnostics.

## Funding Statement

This study was performed as part of a quality assurance initiative at BioExcel Diagnostics. No external funding was received.

Declaration of AI and AI-assisted Technologies in the Writing Process During the preparation of this work, the authors used Grammarly and OpenAI’s GPT-4&5 to enhance clarity and improve formatting. After using these tools, the authors reviewed and edited the content as needed and take full responsibility for the content of the publication.

## Data Availability Statement

All data analyzed in this study were generated as part of routine clinical testing at BioExcel Diagnostics and a commercial CLIA laboratory. De-identified data may be made available upon reasonable request to the corresponding author, subject to institutional policies.

## Supplementary Figures

**Figure S1. Flowchart illustrating the classification of 107 wound infection cases based on conventional diagnostic criteria.** Of the total 107 cases, two were excluded due to mixed or unidentified results, and 12 were deemed inconclusive due to limitations in aerobic or anaerobic culture. The remaining 93 cases were classified into True Positives (n=58), False Positives (n = 9), False Negatives (n=1), and True Negatives (n = 29), False Negatives (n=1), and True Negatives (Fully Concordant (n=17), Partially Concordant (n=25), and cases with Unidentified Additional Organisms (n=16).

**Figure S2. Turnaround time (TAT) comparison between PCR and culture.** Case-wise turnaround times (TATs) for PCR (median 20 h; interquartile range [IQR], 4.5 h) and the reference laboratory’s culture workflow (median 120 h; IQR, 0 h). Wilcoxon signed-rank test, p = 2.57 × 10⁻¹⁹. The reference lab’s fixed mid-distribution TAT (IQR = 0) suggests batch or scheduled reporting; PCR shows modest variability around a ∼20-hour median.

**Figure S3. Receiver operating characteristic (ROC) curve showing the predictive value of pan-bacterial burden (Ct_16_S) for infection status.** AUC = 0.75, with Youden’s optimal threshold at Ct ≤ 18.4 (sensitivity = 78%, specificity = 66%).

## Notes

### Competing Interest Statement

MD is a direct member of BioExcel Diagnostics, LLC
K.H.L & H.L.M are indirect membersof BioExcel Diagnostics, LLC

### Funding Statement

This study did not receive any funding

### Author Declarations

The Ethics Committee/Institutional Review Board (IRB) of BioExcel Diagnostics waived ethical approval for this study, as all work was performed on de-identified, non-trackable data collected as part of routine quality assurance processes at BioExcel Diagnostics.

## References

1. Ba Lipsky, Ar Berendt, Pb Cornia, Jc Pile, Ej Peters, Dg Armstrong, Et Al. 2012. 2012 Infectious Diseases Society of America clinical practice guideline for the diagnosis and treatment of diabetic foot infections. Clin Infect Dis 54:e132–e173.

2. Dg Armstrong, Ajm Boulton, Sa Bus. 2017. Diabetic foot ulcers and their recurrence. N Engl J Med 376:2367–2375.

3. Se Gardner, Ra Frantz, Cl Saltzman, Sl Hillis, H Park, M Scherubel. 2006. The neuropathic diabetic foot ulcer: A study of wound microbiology and bacteremia. J Wound Ostomy Continence Nurs 33:640–647.

4. Dm Citron, Ej Goldstein, Cv Merriam, Ba Lipsky, Ma Abramson. 2007. Bacteriology of moderate-to-severe diabetic foot infections and in vitro activity of antimicrobial agents. J Clin Microbiol 45:2819–2828.

5. Jp Lavigne, A Sotto, C Dunyach-Remy. 2016. New molecular insights into bacterial pathogen identification and antibiotic susceptibility testing in wounds. Adv Wound Care 5:431–439.

6. Lr Kalan, Ma Loesche, Bp Hodkinson, K Heilmann, G Ruthel, Se Gardner, Et Al. 2019. Redefining the chronic-wound microbiome: Fungal communities are prevalent, dynamic, and associated with delayed healing. mBio 10:e00930–19.

7. K Smith, A Collier, Em Townsend, Le O’donnell, Am Bal, J Butcher, Et Al. 2021. Culture versus molecular identification of bacteria in wound samples. Diagn Microbiol Infect Dis 100:115378.

8. Se Dowd, Y Sun, Pr Secor, Dd Rhoads, Bm Wolcott, Ga James, Et Al. 2008. Evaluation of the bacterial diversity in chronic wounds using 16S rDNA pyrosequencing. BMC Microbiol 8:43.

9. M Malone, K Johani, So Jensen, Ib Gosbell, Hg Dickson, H Hu, Et Al. 2017. Next generation DNA sequencing of tissues from infected diabetic foot ulcers. EBioMedicine 21:142–149.

10. Dd Rhoads, Rd Wolcott, Y Sun, Se Dowd. 2012. Comparison of culture and molecular identification of bacteria in chronic wounds. J Clin Microbiol 50:3821–3829.

11. Liu C. M., Et Al. 2013. Bacterial community structure and interactions in a healthy human oral cavity and the effects of mechanical cleaning. mBio 4:e00592–12.

12. Van Smeden M., Naaktgeboren C. A., Reitsma J. B., Moons K. G., De Groot J. A. 2014. Latent class models in diagnostic studies when there is no reference standard--a systematic review. Am J Epidemiol 179:423–31.

13. Se Dowd, Rd Wolcott, Y Sun, T Mckeehan, E Smith, D Rhoads. 2012. Molecular diagnostics and swab techniques reduce false-negative culture results in chronic wound diagnostics. Int J Mol Sci 13:2535–2550.

14. Hansen W. L. J., Et Al. 2013. Real-Time PCR-Based Semi-Quantitative Breakpoint to Aid Diagnosis of Urinary Tract Infections. European Journal of Clinical Microbiology & Infectious Diseases 32:655–661.

15. Rao S. N., Manissero D., Steele V. R., Pareja J. 2020. A Systematic Review of the Clinical Utility of Cycle Threshold Values in the Context of COVID-19. Infect Dis Ther 9:573–586.

16. Chun J. W., Lee B., Park W. S., Han N., Hong E. K., Park E. Y., Han S. S., Park S. J., Kim T. H., Lee W. J., Woo S. M. 2021. RRM1 Expression as a Prognostic Biomarker for Unresectable or Recurrent Biliary Tract Cancer Treated with Gemcitabine plus Cisplatin. J Clin Med 10:4652.

17. Jh Melendez, Ym Frankel, At An, L Williams, Lb Price, Ny Wang, Et Al. 2010. Real-time PCR assays compared to culture-based approaches for identification of bacteria in chronic wound infections. J Clin Microbiol 48:3953–3959.

18. Barnaud G., Arlet G., Verdet C., Gaillot O., Lagrange P. H., Philippon A. 1998. Salmonella enteritidis: AmpC plasmid-mediated inducible beta-lactamase (DHA-1) with an ampR gene from Morganella morganii. Antimicrob Agents Chemother 42:2352–8.

19. Jacoby G. A., Munoz-Price L. S. 2005. The new beta-lactamases. N Engl J Med 352:380–91.

20. Naas Thierry, Coignard Bruno, Carbonne Anne, Blanckaert Karen, Bajolet Olivier, Bernet Christine, Bereksi Najet, Ducellier Dominique, Nordmann Patrice. 2005. Emergence of OXA-23-producing carbapenem-resistant Acinetobacter baumannii in a tertiary care hospital. Journal of Clinical Microbiology 43:4826–4829.

21. Kl Frank, Jl Del Pozo, R Patel. 2008. From clinical microbiology to infection pathogenesis: how daring to be different works for Staphylococcus lugdunensis. Clin Microbiol Rev 21:111–133.

22. Gs Collins, Jb Reitsma, Dg Altman, Kgm Moons. 2015. Transparent reporting of a multivariable prediction model for individual prognosis or diagnosis (TRIPOD): explanation and elaboration. Ann Intern Med 162:W1–73.

23. A Niculescu-Mizil, R Caruana. Predicting good probabilities with supervised learning, p 625–32. *In* (ed),

24. Li J., Jiang B., Fine J. P. 2013. Multicategory reclassification statistics for assessing improvements in diagnostic accuracy. Biostatistics 14:382–94.

25. Behera Himanshu Sekhar, Chayani Nirupama, Bal Madhusmita, Khuntia Hemant Kumar, Pati Sanghamitra, Das Sashibhusan, Ranjit Manoranjan. 2021. Identification of population of bacteria from culture negative surgical site infection patients using molecular tool. BMC Surgery 21:28.

26. Gramberg Meryl Cinzía Tila Tamara, Knippers Carmen, Lagrand Rimke Sabine, Van Hattem Jarne Marijn, De Goffau Marcus Christofoor, Budding Budding Andries Edward, Davids Mark, Matamoros Sebastien, Nieuwdorp Max, De Groot Vincent, Heijer Martin Den, Sabelis Louise Willy Elizabeth, Peters Edgar Josephus Gerardus. 2023. Concordance between culture, Molecular Culture and Illumina 16S rRNA gene amplicon sequencing of bone and ulcer bed biopsies in people with diabetic foot osteomyelitis. BMC Infectious Diseases 23:505.

27. Lieu A., Mah J., Peirano G., Somayaji R., Church D. 2022. Microbiological Characterization of Actinotignum schaalii Strains Causing Invasive Infections during a Multiyear Period in a Large Canadian Health Care Region. Microbiol Spectr 10:e0344222.

28. Delgado-Viscogliosi Pascale, Regnault Bertrand, Madelaine Claire, Rodier Marie-Hélène. 2009. Development and evaluation of viability PCR for Legionella pneumophila in water samples. Applied and Environmental Microbiology 75:3192–3200.

29. Weese J. S., Saab M., Moore A., Cai H., Mcclure J. T. 2023. Relationship between quantitative real-time PCR cycle threshold and culture for detection of Streptococcus equi subspecies equi. Can Vet J 64:549–552.

30. Karam Nadine, Martinez Ana, Clarke Thomas, Smith Edward. 2024. Diagnosing gastrointestinal infections based on cycle threshold cut-offs of PCR assays. Microbiology Spectrum 12:e01234–24.

31. Von Hertwig Clara, Gonzalez Luis, Perez Maria, Johnson Robert. 2024. Quantification of viable Salmonella in low-moisture foods by PMA-qPCR and culture. Food Microbiology 45:120–130.

32. Hamza Omar, Farouq Mohamed, Saeed Yasmin, El-Mahdy Heba. 2024. Combining Gram Stain and Quantitative 16S rRNA PCR for Enhanced Diagnosis of Hospital-Acquired Pneumonia. Journal of Clinical Microbiology 62:e01423–23.

33. Limmathurotsakul Direk, Wuthiekanun Vanaporn, Day Nicholas P. J., Peacock Sharon J. 2010. Improved detection of Bacillus species in blood culture through use of Bayesian latent class modeling. PLoS ONE 5:e12485.

34. Melendez J. H., Rojas N. L., Miller C., Chappel A. R., Cavuoti D., Hospenthal D. R., Murray C. K., Akers K. S. 2010. Molecular versus traditional techniques for identification of bacteria in chronic wounds. Clinical Microbiology and Infection 16:1762–1769.

35. Cederlöf Ingela, Toft Nils, Aalbæk Bent, Klaas Ilka C., Eriksen Nina H., Nielsen S. S. 2012. Latent class analysis of diagnostic characteristics of PCR and bacteriological culture for detection of Staphylococcus aureus in milk. Acta Veterinaria Scandinavica 54:65.

36. Schumacher S. G., Van Smeden M., Dendukuri N., Joseph L., Nicol M. P., Pai M., Zar H. J. 2016. Diagnostic Test Accuracy in Childhood Pulmonary Tuberculosis: A Bayesian Latent Class Analysis. Am J Epidemiol 184:690–700.

37. Le T. H., Nguyen T. Q., Trinh M. H., Pham H. N., Dang T. M., Hoang T. M. 2020. Diagnostic accuracy of smear, culture, and Xpert MTB/RIF in tuberculous meningitis: A latent class analysis. Journal of Infection in Developing Countries 14:817–823.

38. Altmann M., Walker T., Swoboda S., Lipsky B. A. 2025. Diagnostic utility of Gram stain and culture in diabetic foot infection: A comparative study. Journal of Clinical Medicine 14:145.

39. Branch-Elliman W., Sturgeon D., Karchmer A. W., Mull H. J. 2021. Association Between Diabetic Foot Infection Wound Culture Positivity and 1-Year Admission for Invasive Infection: A Multicenter Cohort Study. Open Forum Infect Dis 8:ofab172.

40. Rhoads Daniel D., Wolcott Randall D., Sun Yan, Dowd Scot E. 2012. Comparison of Culture and Molecular Identification of Bacteria in Chronic Wounds. International Journal of Molecular Sciences 13:2535–2550.

41. Cummings Lisa A., Hoogestraat Daniel R., Rassoulian-Barrett Sara L., Rosenthal Christopher A., Salipante Stephen J., Cookson Brad T., Hoffman Noah G. 2020. Comprehensive evaluation of complex polymicrobial specimens using next generation sequencing and standard microbiological culture. Scientific Reports 10:5446.

42. Gardner Sue E., Hillis Stephen L., Heilmann Kris, Segre Julia A., Grice Elizabeth A. 2013. The Neuropathic Diabetic Foot Ulcer Microbiome Is Associated With Clinical Factors. Diabetes 62:923–930.

43. Hung S. Y., Chiu C. H., Huang C. H., Lin C. W., Yeh J. T., Yang H. M., Huang Y. Y. 2022. Impact of wound microbiology on limb preservation in patients with diabetic foot infection. J Diabetes Investig 13:336–343.

44. Abayasekara Lalanika M., Perera Jennifer, Chandrasekharan Vishvanath, Gnanam Vaz S., Udunuwara Nisala A., Liyanage Dileepa S., Bulathsinhala Nuwani E., Adikary Subhashanie, Aluthmuhandiram Janith V. S., Thanaseelan Chrishanthi S., Tharmakulasingam D. Portia, Karunakaran Tharaga, Ilango Janahan. 2017. Detection of bacterial pathogens from clinical specimens using conventional microbial culture and 16S metagenomics: a comparative study. BMC Infectious Diseases 17:631.

